# Invasive Mechanical Ventilation Duration Prediction using Survival Analysis

**DOI:** 10.1101/2022.12.15.22283535

**Authors:** Yawo Kobara, Felipe F. Rodrigues, Camila P. E. de Souza, Megan Wismer

## Abstract

Invasive mechanical ventilation is one of the leading life support machines in the intensive care unit (ICU). By identifying the predictors of ventilation time upon arrival, important information can be gathered to improve decisions regarding capacity planning.

In this study, first-day ventilated patients’ ventilation time was analyzed using survival analysis. The probabilistic behaviour of ventilation time duration was analyzed and the predictors of ventilation time duration were determined based on available first-day covariates.

A retrospective analysis of ICU ventilation time in Ontario was performed with data from ICU patients obtained from the Critical Care Information System (CCIS) in Ontario between July 2015 and December 2016. As part of the protocol for inclusion, a patient must have been connected to an invasive ventilator upon arrival to the ICU. Parametric survival methods were used to characterize ventilation time and to determine associated covariates. Parametric and non-parametric methods were used to determine predictors of ventilation duration for first-day ventilated patients.

A total of 128,030 patients visited the ICUs between July 2015 and December 2016. 51,966 (40.59%) patients received invasive mechanical ventilation on arrival. Analysis of ventilation duration suggested that the log-normal distribution provided the best fit to ventilation time, whereas the log-logistic Accelerated Failure Time model best describes the association between the covariates and ventilation duration. ICU site, admission source, admission diagnosis, scheduled admission, scheduled surgery, referring physician, central venous line treatment, arterial line treatment, intracranial pressure monitor treatment, extra-corporeal membrane oxygen treatment, intraaortic balloon pump treatment, other interventions, age group, pre-ICU LOS, and MODS score were significant predictors of the ICU ventilation time.

The results show substantial variability in ICU ventilation duration for different ICUs, patient’s demographics, and underlying conditions, and highlight mechanical ventilation as an important driver of ICU costs.

The predictive performance of the proposed model showed that both the model and the data can be used to predict an individual patient’s ventilation time and to provide insight into predictors.

## 1. Introduction

Mechanical ventilation (MV) is defined as the use of a breathing support machine that takes over the breathing process in patients who cannot breathe properly on their own. MV is one of the life support alternatives provided by the Intensive Care Unit (ICU) that differentiate it from other hospital units. There are two forms of MV: invasive and non-invasive. Non-invasive ventilation (NIV) is the delivery of oxygen through a face mask. According to Hyzy and McSparron [2022], invasive mechanical ventilation (IMV) is “the delivery of positive pressure to the lungs through an endotracheal or tracheostomy tube.” During IMV, a ventilation machine (also called a ventilator) forces a predetermined mixture of air (i.e., oxygen and other gases) into the central airways that then flows into the alveoli [Hyzy and McSparron, 2022, Luo et al., 2017]. A patient may need a ventilator when there is a low oxygen level in the blood or severe shortness of breath from an infection such as pneumonia, SARS or COVID-19. Over 20 million patients worldwide per year are treated with mechanical ventilation [Urner et al., 2020, Adhikari et al., 2010].

Since the outbreak of the COVID-19 pandemic, one of the most globally cited causes for the inability of ICUs to manage patients appropriately has been the lack of ventilators [Beitler et al., 2020, Elegant, 2020, Pearce, 2020, Begley, 2020, Iyengar et al., 2020]. However, the concern about an insufficient supply of ICU beds and ventilators to handle critically ill patients is old. COVID-19 sparked a debate on when and how ventilators should be used within the ICU. Before COVID-19, there have been very few publications that modelled the use, demand, and practice of ventilation on ICU patients. Kacmarek [2011] gave a detailed history of the evolution of these human breathing aids in medicine. Ventilators are not built into ICU beds like other organ support machines. Nevertheless, as many as 90% of ICU patients require ventilation [Pearce, 2020]. Previous papers in the literature associate ICU ventilator use with ICU bed use, but not all patients in the ICU use ventilators, and their use is patient-state-dependent.

From a managerial perspective, it is important to know the distribution of the time that patients are reliant upon IMV to predict ventilation demand. In this case, survival models are often used to determine when to stop invasive ventilation. Survival analysis is extensively used in health care but the application of such models to ventilation time is not common. To the authors’ knowledge, no study has analyzed ventilation time using survival models.

Most studies on ventilation time in the literature were interested in classifying the duration of mechanical ventilation into two categories: prolonged versus short [Troche and Moine, 1997, Estenssoro et al., 2005, Dimopoulou et al., 2003, Abujaber et al., 2020, Figueroa-Casas et al., 2015, Trouillet et al., 2009, Aung et al., 2018]. Prolonged mechanical ventilation (PMV) was defined differently within each study. Estenssoro et al. [2005] described it as being mechanical ventilation for longer than 21 days, Dimopoulou et al. [2003] defined PMV as longer than seven days, Trouillet et al. [2009] defined it as ventilation for longer than three days, and Lésegaréet al. [2001] described it as ventilation for more than one day. Logistic regression, linear regression and machine learning methods are the primary tools used in the literature to model ventilation time and identify significant predictors of PMV.

Dimopoulou et al. [2003] investigated PMV in patients with blunt thoracic trauma and found that advancing age, severity of head injuries, and bilateral thoracic injuries were significant in predicting PMV. Trouillet et al. [2009] gathered data on patients undergoing cardiac surgery. They found that a post-operative score could be used to identify patients eligible for rapid weaning of ventilation on day three, which reduced the need for PMV. Lésegaré et al. [2001] took a group of coronary artery bypass graft patients, identified the predictors of PMV, and found that intraoperative complications significantly impacted those patients who required prolonged mechanical ventilation. Trouillet et al. [2011] investigated the outcomes of two groups of severely ill patients who required mechanical ventilation. One group received early percutaneous tracheotomy and the other received prolonged intubation. When the two treatments, it was found that early tracheotomy provided no benefit in terms of mortality rate or length of stay. Esteban et al. [2002] found that factors at the start of mechanical ventilation and complications of critical illness influenced the outcome of patients receiving mechanical ventilation.

Logistic regression is limited in predicting the duration of mechanical ventilation, aside from classifying it. Moreover, when studies used linear regression, the results were often unreliable. Seneff et al. [1996] analyzed an individual patient’s duration of mechanical ventilation using linear regression and found an *R*^2^ of 0.18, which indicates that the predictions were not reliable. Aung et al. [2018] used multiple linear regression to identify variables independently associated with the duration of mechanical ventilation duration and obtained an *R*^2^ of 0.235.

Abujaber et al. [2020] and Sayed et al. [2021] used machine learning models but could not find direct relationships between the predictors and the mechanical ventilation duration. Abujaber et al. [2020] started with logistic regression and built upon this by creating Artificial Neural Networks, Support Vector Machines, Random Forests, and Decision Trees to predict prolonged mechanical ventilation. Ultimately, Support Vector Machines gave the best prediction in this study, with an accuracy of 0.79, but had the limitation of being able to find only the predictor importance. Sayed et al. [2021] attempted to predict the duration of mechanical ventilation using machine learning models. When using the Light Gradient Boosting Machine, it was found that predictors gathered before the third ICU day could be used, enabling mechanical ventilation to be predicted earlier than other machine learning models such as random forest and extreme gradient boosting.

This study predicted ventilation duration in the ICU using patient information available on arrival. The distribution of ventilation duration was identified and the existing differences in terms of first-day treatments, ICU sites, admission source, referring physician, patient category, sex, NEMS, and MODS were evaluated. This study demonstrates the importance of patient information available at arrival when predicting ventilation duration among ICU patients. Hence, these results are important in ventilation planning and management.

## 2. Methods

### 2.1 Study Design and Data Collection

This is a retrospective study designed to predict the time distribution of ICU patients that were connected to IMV on arrival using information available on arrivals, such as Demographics, First-day treatments, NEMS and MODS scores. The objective was to determine the predictors of ventilation time after the first day and predict ventilation duration. Survival analysis was performed using a large dataset of variables obtained from the Critical Care Information System (CCIS) Ontario database.

The CCIS dataset contains information from July 2015 to December 2016. The data includes forty-five variables. Priestap et al. [2020] used the CCIS dataset to predict ICU mortality and provide detailed information on the data collection procedure, the variables and the ICUs in the CCIS database. The following subset of covariates on patient arrival was used in this study: *Basic Monitoring, Central Venous Line, Arterial Line, Intracranial Pressure Monitor, Dialysis, Extra-corporeal Membrane Oxygen, Intra-aortic Balloon Pump, Other Interventions Within this Unit, Interventions Outside this Unit, the nursing workload proxy by the First-day NEMS score, demographic information (Age, Sex), and the MODS Score*. For external model validation, data from London Health Science Center (LHSC) were also used in this study. The LHSC datasets contain information from January 2020 to May 2021.

The data were pre-processed to remove duplicates, transform variables and create new variables needed for the analysis. The data contain records of 128,030 patients admitted into the ICUs. 40.59% (51,966) of those patients received IMV on arrival. Table 1 presents the summary of the patients’ ventilation status on arrival. Of the patients that received IMV upon arrival, those with missing information and were excluded to obtain 49,703 (i.e., 38.82% of total ICU patients). Furthermore, those with ventilation time greater than 60 days were excluded and the 49,467 (99.53 % of patients connected on arrival) remaining were used in the study. Table 2 shows the sex and censoring distribution of patient information used in this research. Censored patients include those discharged to the Complex Continuing Care Facility, other hospitals, the Level 3 Unit, and Outside the ICU while on a ventilator.

**Table 1:**
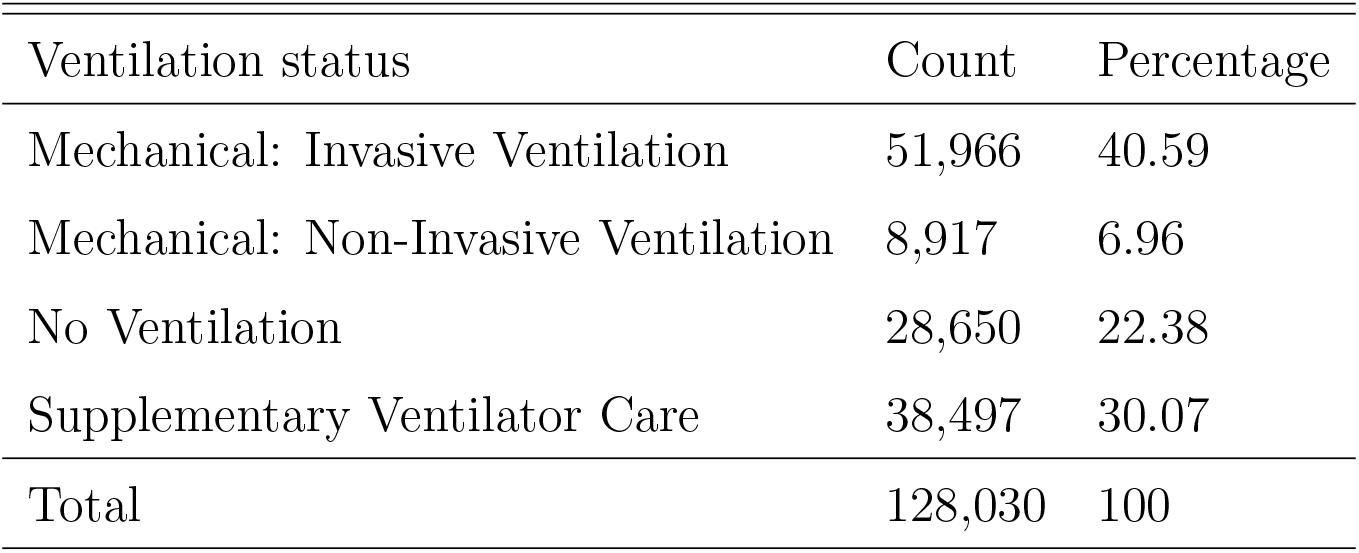
First-day ventilation frequency

**Table 2:**
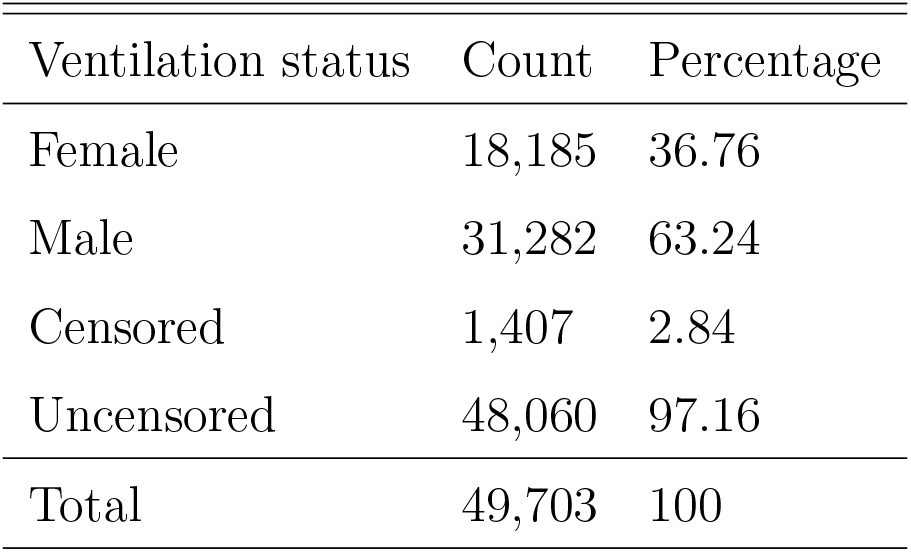
Gender and censor status distribution of data used

| Ventilation status | Count | Percentage |
| --- | --- | --- |
| Female | 18,185 | 36.76 |
| Male | 31,282 | 63.24 |
| Censored | 1,407 | 2.84 |
| Uncensored | 48,060 | 97.16 |
| Total | 49,703 | 100 |

This study was approved by the Research Ethics Review Committee at King’s University College.

### 2.2 Statistical analysis

#### Descriptive analysis

Descriptive statistical analysis was performed, reporting the following measures for continuous variables: mean, standard deviation, skewness, kurtosis, and quartiles. For modelling, the continuous variables were grouped into categories. Categorical variables were reported as counts and percentages.

#### Non-parametric survival analysis

Kaplan-Meier analysis was used to construct non-parametric survival curves based on patients’ age and sex. The log-rank (Mantel-Cox) test and the Kruskal-Wallis test were used for multiple comparisons between sub-groups.

#### Parametric survival analysis

Various classical distributions were fitted to the observed ventilation time to identify the probability distribution function (pdf) with the best fit. To assess goodness-of-fit, P-P plots, as well as three regularly used goodness-of-fit tests: Kolmogorov-Smirnov, Anderson-Darling, and ChiSquared were used. Appropriate maximum likelihood estimates of the parameters were obtained with their respective 95% confidence limits based on the probability distribution function. Parametric Accelerated Failure Time (AFT) modelling was done by randomly dividing the dataset into a “training” and a “testing” set (a training set with 70% of the observations and a testing set with 30%). The training set was used for modelling, and the test set was used for model performance prediction. External data for different years gathered from the London Health Science Center (LHSC) were used to validate the model.

All covariates included in the analysis were obtained on arrival and are included based on their availability, clinical relevance, statistical significance, and possible association with ICU LOS or mortality in the literature. This study followed the variable selection approach as outlined in Collett [2015]. This approach fits a univariate model for each covariate, identifying significant predictors after which a multivariate model was fitted with all significant univariate predictors, eliminating insignificant variables using backwards selection. Graphical methods, the likelihood ratio, AIC and BIC criteria were used to compare and select the AFT models (Exponential, Weibull, Log-normal, and Log-logistic). The validity of the model was ascertained using external data. All tests presented are two-sided, and a *p*-value < 0.05 is considered significant.

#### Predictive performance

To assess predictive performance, the best fitted model was applied to the test set and considered the following metrics were considered: Mean Squared Error (MSE), Mean Absolute Error (MAE), Percent bias (PBIAS), and Nash-Sutcliffe efficiency (NES). These metrics were calculated as follows.

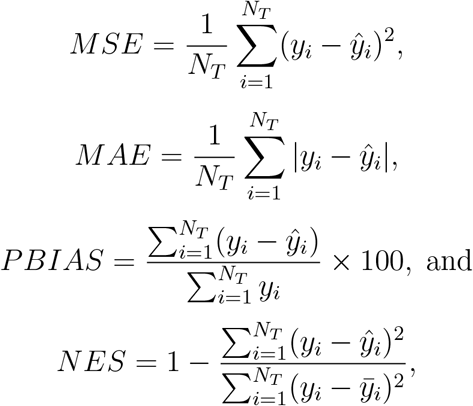

where *N*_*T*_ is the total number of observations in the test set and, *y*_*i*_ and *ŷ*_*i*_ are respectively the observed and predicted time on ventilation for the *i*th observation in the test set.

#### Statistical software

Analyses were performed using R version 4.1.2. Statistical modelling used the *glmnet, flexsurv, SurvRegCensCov, survival*, and *surminer* packages.

## 3. Results

### 3.1 Descriptive Analysis

Table 3 summarizes the number and proportion of patients that received each of the various treatments at ICU arrival. The most common were basic monitoring (99.93 % of patients), an arterial line (81.76 %), and a central venous line (72.95 %). Variation in treatment patterns showed that 98.34% had no intracranial pressure monitoring, 97.13% had no dialysis, 99.73% had no extracorporeal membrane oxygen, 98.59% had no intra aortic balloon pump 67.38 % had no other interventions within this unit and 78.15% had no interventions outside this unit.

**Table 3:**
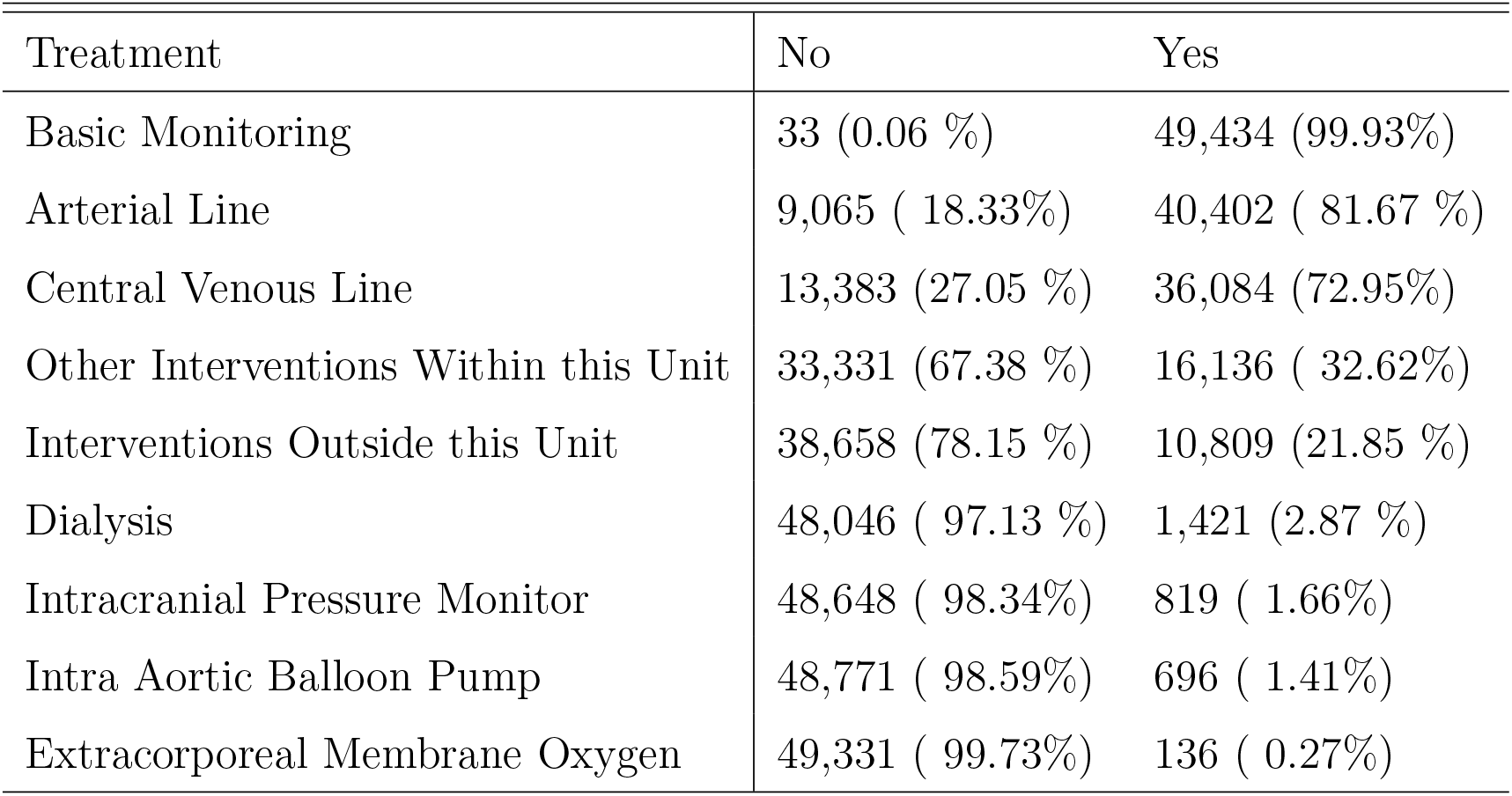
Distribution of treatments IMV patients received

| Treatment | No | Yes |
| --- | --- | --- |
| Basic Monitoring | 33 (0.06 %) | 49,434 (99.93%) |
| Arterial Line | 9,065 ( 18.33%) | 40,402 ( 81.67 %) |
| Central Venous Line | 13,383 (27.05 %) | 36,084 (72.95%) |
| Other Interventions Within this Unit | 33,331 (67.38 %) | 16,136 ( 32.62%) |
| Interventions Outside this Unit | 38,658 (78.15 %) | 10,809 (21.85 %) |
| Dialysis | 48,046 ( 97.13 %) | 1,421 (2.87 %) |
| Intracranial Pressure Monitor | 48,648 ( 98.34%) | 819 ( 1.66%) |
| Intra Aortic Balloon Pump | 48,771 ( 98.59%) | 696 ( 1.41%) |
| Extracorporeal Membrane Oxygen | 49,331 ( 99.73%) | 136 ( 0.27%) |

**Table 4:** Selected survival estimates from the K-M curve.

| t | n.risk | n.event | n.censor | surv (95% CI) |
| --- | --- | --- | --- | --- |
| 1 | 49,467 | 15,125 | 646 | 0.694 (0.698, 0.690) |
| 7 | 9,100 | 1,270 | 28 | 0.167 (0.170, 0.163) |
| 10 | 5,879 | 698 | 20 | 0.112 (0.115, 0.109) |
| 30 | 791 | 68 | 2 | 0.016 (0.018, 0.015) |
| 50 | 119 | 12 | 0 | 0.003 (0.003, 0.002) |
| 55 | 56 | 12 | 0 | 0.001 (0.001, 0.001) |

n.risk, n.event, n.censor, and surv are number at risk, number of events, number censored, and Survival probability at time $t$ .

Figure 3.1 shows a bar plot of the admission sources, admitting diagnosis, referring physician specialty, and patient category. The admission sources included the ward (5,607, 11.33%), downstream unit (ie. Level 2 and Level 3 units) (2,971, 6.00%), the emergency department (ED) (12,507, 25.28%), home (191, 0.39%), hospital outside and within the province(5,210, 10.53%), the operating room (OR) (22,608, 45.70%), and other sources (373, 0.75%). The other sources included complex continuing care facilities, rehabilitation centers, outside the province, and others.

ICU admitting diagnoses were categorized as Cardiovascular (22,269, 45.00%), Gastrointestinal (2,920, 5.90%), Neurological(4,652, 9.40), Trauma (1,927, 3.90%), and Other (17699, 35.78). Other diagnoses included patients with the following categories of disease: Genitourinary, Metabolic, Endocrine, Musculoskeletal, Skin, Oncology, Haematology and Others. Referring physician specialties were grouped into medical (16930, 34.22%), respiratory (1,364, 2.76%), surgical (21,848, 44.17%), and other (9,325, 18.85%). Other referring physician specialist includes Dermatology, Psychiatry, Oncology, Haematology, Ophthalmology, Orthopaedic, and others.

[!h] [ICU admission source (other source includes Complex Continuing Care Facility - Within and Outside, Inpatient - Rehab, Outside province, Rehab Facility - Within and Outside, and Other - Outside and Within))] [ICU admission diagnosis (Other diagnosis includes patients with the following categories of disease: Genitourinary, Metabolic, Endocrine, Musculoskeletal, Skin, Oncology / Haematology and Other.)] [Referring physician specialty. (Other referring physician specialties included: Dermatology, Psychiatry, Oncology, Haematology, Ophthalmology, Orthopaedic, and other.)]

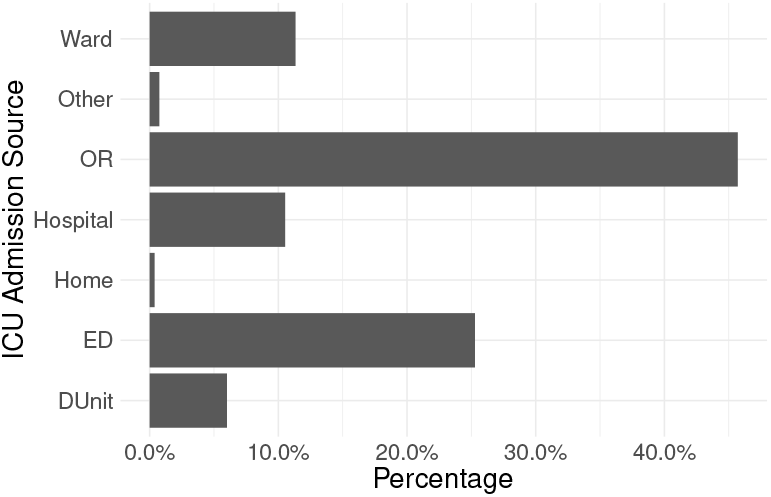

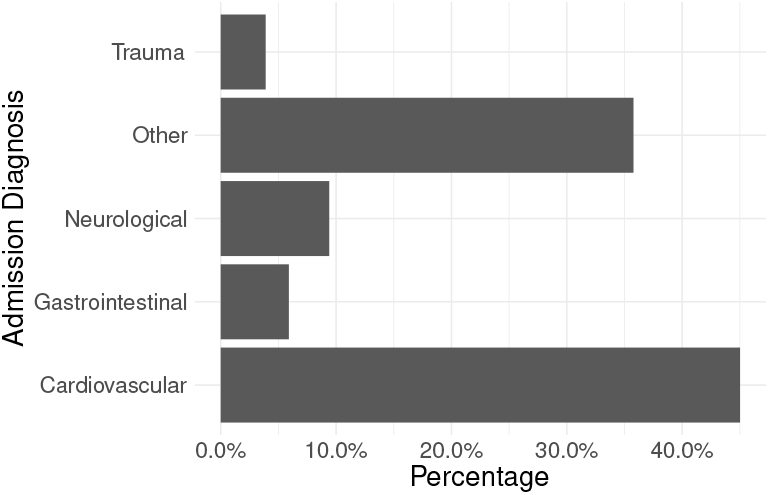

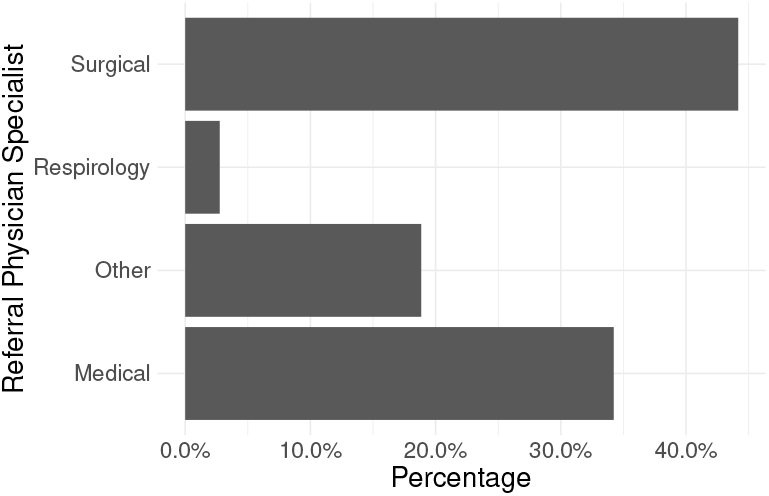

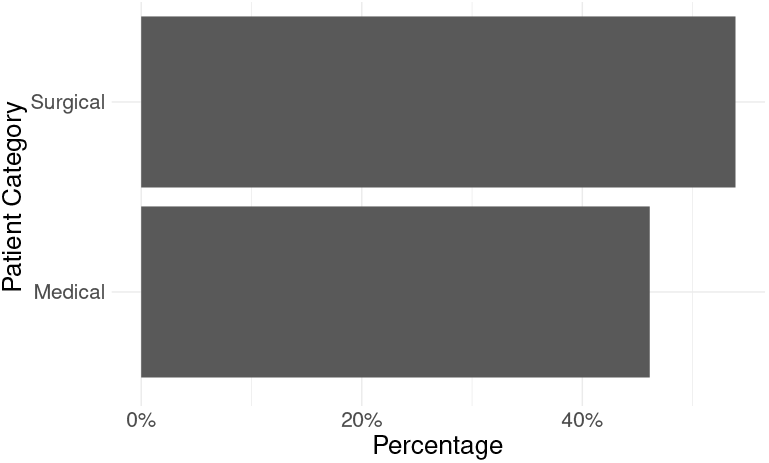

[Patient category]

Table 6 provides descriptive statistics for the continuous variables., and Table 5 provides descriptive statistics for ventilation time under various patient categories. On average, ventilation time was 4.57 (sd = 6.57) days, the NEMS score was 29.09 (sd = 6.85), and the MODS score was 5.57 (sd = 3). Table 14 tabulates the baseline characteristics of the number of events under each category and the results of the Log-rank test, which compares the differences in ventilation times between the independent groups on each covariate. For ease of readability, the results of the Log-rank test are shown in Table 7.

**Table 5:** Descriptive statistics of ventilation time under various patient categories.

| Variables | | Count | Mean | sd | Skew | Kurt | $Q_{0.25}$ | $Q_{0.5}$ | $Q_{0.75}$ |
| --- | --- | --- | --- | --- | --- | --- | --- | --- | --- |
| Vent days |  | 49467 | 4.57 | 6.57 | 3.65 | 17.02 | 1 | 2 | 5 |
| (Sex) | Female | 18185 | 4.84 | 6.82 | 3.59 | 16.35 | 1 | 2 | 5 |
|  | Male | 31282 | 4.41 | 6.41 | 3.69 | 17.39 | 1 | 2 | 4 |
| (NEMS) | 0 – 22 | 629 | 4.79 | 7.21 | 3.75 | 17.09 | 1 | 2 | 5 |
|  | 23 – 29 | 11823 | 4.03 | 6.06 | 4.05 | 21.11 | 1 | 2 | 4 |
| | $\geq 30$ | 37015 | 4.74 | 6.70 | 3.55 | 16.00 | 1 | 2 | 5 |
| (Age group) | 0 – 39 | 4355 | 4.91 | 6.47 | 3.30 | 14.18 | 2 | 2 | 5 |
|  | 40 – 79 | 38009 | 4.54 | 6.61 | 3.67 | 17.06 | 1 | 2 | 5 |
| | $\geq 80$ | 7103 | 4.51 | 6.36 | 3.77 | 18.65 | 1 | 2 | 5 |
| (MODS) | 1 | 4389 | 2.80 | 4.66 | 5.44 | 39.28 | 1 | 1 | 2 |
|  | 1 – 4 | 14185 | 4.02 | 6.09 | 4.22 | 23.00 | 1 | 2 | 4 |
|  | 5 – 8 | 16070 | 4.89 | 6.75 | 3.40 | 14.63 | 1 | 2 | 5 |
|  | 9 – 12 | 6861 | 5.94 | 7.73 | 3.00 | 11.10 | 2 | 3 | 7 |
| | $\geq 13$ | 7962 | 4.71 | 6.52 | 3.55 | 16.28 | 1 | 2 | 5 |
| Source | Downstream | 2971 | 6.93 | 8.16 | 2.57 | 8.03 | 2 | 4 | 8 |
|  | ED | 12507 | 5.50 | 6.78 | 3.24 | 13.79 | 2 | 3 | 6 |
|  | Home | 191 | 5.75 | 6.07 | 2.06 | 4.38 | 2 | 3 | 7 |
|  | Hospital | 5210 | 6.81 | 8.08 | 2.83 | 9.86 | 2 | 4 | 8 |
|  | OR | 22608 | 2.68 | 4.36 | 5.91 | 46.25 | 1 | 2 | 2 |
|  | Other | 373 | 7.20 | 8.52 | 2.65 | 8.15 | 2 | 4 | 9 |
|  | Ward | 5607 | 6.54 | 8.40 | 2.80 | 9.32 | 2 | 3 | 8 |
| Diagnosis | Cardiovascular | 22269 | 2.87 | 4.77 | 5.61 | 41.35 | 1 | 2 | 2 |
|  | Gastrointestinal | 2920 | 4.76 | 5.97 | 3.30 | 13.99 | 2 | 3 | 5 |
|  | Neurological | 4652 | 5.47 | 6.61 | 2.96 | 11.55 | 2 | 3 | 6 |
|  | Other | 17699 | 6.24 | 7.88 | 2.98 | 10.80 | 2 | 3 | 7 |
|  | Trauma | 1927 | 6.34 | 7.13 | 2.54 | 8.77 | 2 | 4 | 8 |
| Referring | Medical | 16930 | 5.91 | 7.33 | 3.09 | 11.94 | 2 | 3 | 7 |
|  | Other | 9325 | 5.59 | 7.24 | 3.18 | 12.86 | 2 | 3 | 6 |
|  | Respirology | 1364 | 7.94 | 9.72 | 2.66 | 8.00 | 2 | 4 | 9 |
|  | Surgical | 21848 | 2.89 | 4.74 | 5.22 | 35.65 | 1 | 2 | 2 |
| Pat Category | Medical | 22811 | 6.10 | 7.60 | 3.05 | 11.58 | 2 | 3 | 7 |
|  | Surgical | 26656 | 3.26 | 5.19 | 4.68 | 28.73 | 1 | 2 | 3 |

**Table 6:** Descriptive statistics of continuous variable

| Variables | Count | Mean | sd | Skew | Kurt | $Q_{0.25}$ | $Q_{0.5}$ | $Q_{0.75}$ |
| --- | --- | --- | --- | --- | --- | --- | --- | --- |
| Day 1 NEMS | 49467 | 35.22 | 6.69 | 0.36 | -0.36 | 28 | 34 | 39 |
| 0 – 22 | 629 | 20.93 | 0.61 | -11.6 | 153.35 | 21 | 21 | 21 |
| 23 – 29 | 11823 | 27.01 | 0.19 | -0.41 | 64.71 | 27 | 27 | 27 |
| $\geq 30$ | 37015 | 38.08 | 5.16 | 0.86 | 0.07 | 34 | 38 | 40 |
| Age | 49467 | 63.84 | 15.88 | -0.70 | 0.20 | 55.25 | 66.05 | 75.29 |
| 18 – 39 | 4355 | 29.19 | 5.95 | -0.10 | -1.19 | 24.14 | 29.47 | 34.38 |
| 40 – 79 | 38009 | 63.85 | 10.00 | -0.42 | -0.64 | 56.75 | 65.00 | 71.96 |
| $\geq 80$ | 7103 | 85.02 | 3.79 | 0.93 | 0.57 | 81.95 | 84.23 | 87.32 |
| Pre ICU LOS | 49467 | 9.05 | 102.61 | 25.76 | 845.44 | 0 | 0 | 2 |
| $\leq 1$ | 35911 | 0.19 | 0.40 | 1.54 | 0.36 | 0 | 0 | 0 |
| 2 – 7 | 7661 | 3.85 | 1.71 | 0.48 | -1.07 | 2 | 4 | 5 |
| $> 7$ | 5895 | 69.78 | 290.10 | 8.93 | 100.69 | 10 | 15 | 29 |
| MODS Score | 49467 | 5.57 | 3 | 0.42 | 0.24 | 4 | 5 | 7 |
| 1 | 4389 | 0.58 | 0.49 | -0.33 | -1.89 | 0 | 1 | 1 |
| 1 – 4 | 14185 | 3.23 | 0.81 | -0.43 | -1.33 | 3 | 3 | 4 |
| 5 – 8 | 16070 | 6.87 | 0.80 | 0.23 | -1.41 | 6 | 7 | 8 |
| 9 – 12 | 6861 | 9.95 | 1.01 | 0.67 | -0.75 | 9 | 10 | 11 |
| $\geq 13$ | 7962 | 6.06 | 2.95 | 2.51 | 4.62 | 5 | 5 | 5 |

**Table 7:** Log-rank test of equality between groups in covariates

| Covariate | $\chi^2$ | df | $p$-value |
| --- | --- | --- | --- |
| Pre ICU LOS | 262 | 2 | <2e-16 |
| Age group | 35 | 2 | 3e-08 |
| Sex | 77.8 | 1 | <2e-16 |
| ICU Site | 3350 | 65 | <2e-16 |
| Admission source | 6875 | 6 | <2e-16 |
| Diagnosis | 5213 | 4 | <2e-16 |
| Referring physician | 5171 | 3 | <2e-16 |
| Patient Category | 3929 | 1 | <2e-16 |
| MODS | 1107 | 4 | <2e-16 |
| NEMS | 157 | 2 | <2e-16 |
| Scheduled admission | 9579 | 1 | <2e-16 |
| Scheduled surgery | 8809 | 1 | <2e-16 |
| Basic Monitoring | 0.1 | 1 | 0.8 |
| Central Venous Line | 53.9 | 1 | 2e-13 |
| Arterial Line | 79.4 | 1 | <2e-16 |
| Intracranial Pressure Monitor | 227 | 1 | <2e-16 |
| Dialysis. | 194 | 1 | <2e-16 |
| Extra corporeal Membrane Oxygen | 79.7 | 1 | <2e-16 |
| Intra Aortic Balloon Pump | 22.5 | 1 | 2e-06 |
| Other Interventions Within this Unit | 878 | 1 | <2e-16 |
| Interventions Outside this Unit | 554 | 1 | <2e-16 |

### 3.2 Non-parametric Analysis

As a preliminary exploration, a non-parametric analysis was conducted of the entire dataset using the Kaplan-Meier method. Figure 3.2 shows the Kaplan-Meier (KM) curve of ICU ventilation time. The KM curve shows the unconditional probability that a subject will experience the event beyond time *t* but does not indicate the proportion of subjects surviving to time *t*. In the present case, survival means becoming independent of ventilation by time *t*.

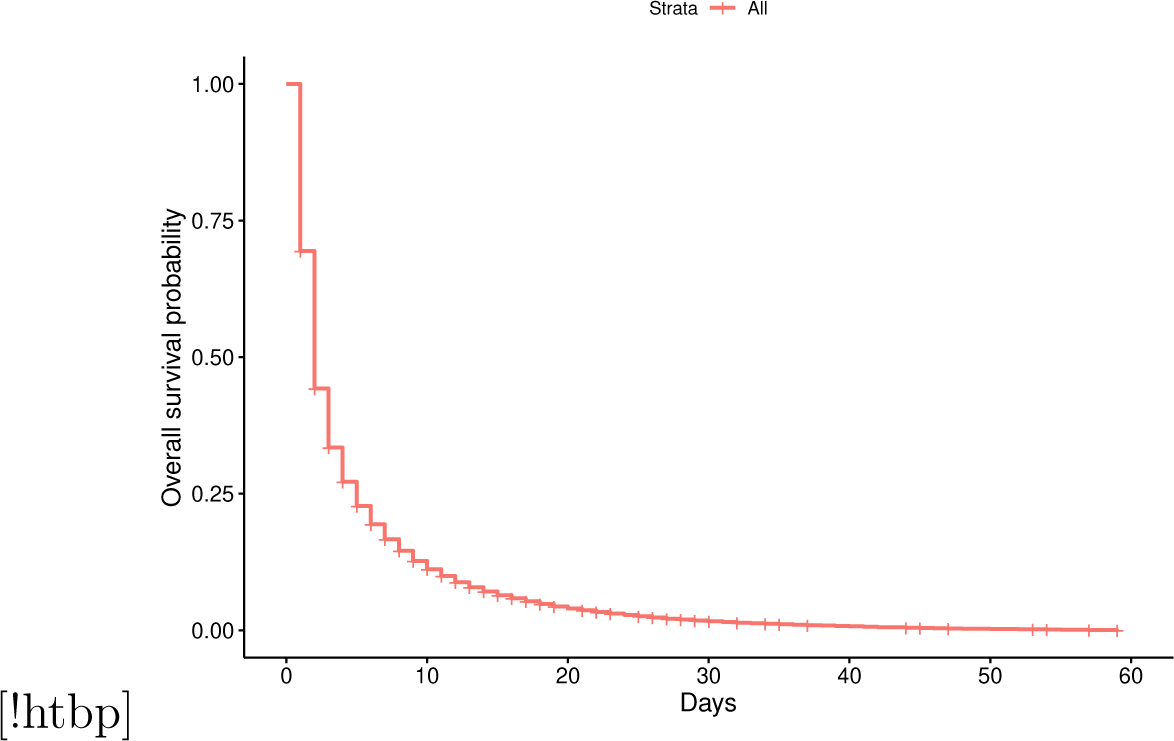

Table 4 tabulates selected KM survival probabilities at specific times and their associated confidence intervals. From Figure 3.2 and Table 4, the probability that a person will remain connected to the IMV longer than one day is approximately 0.69. In other words, about 30% get disconnected from mechanical ventilation after the first day. The probability of patients remaining connected beyond 10 days is about 0.112, indicating that about 90% of ICU patients get disconnected after ten days of IMV.

The patients included 18,185 (36.76 %) females with a mean ventilation time of 4.84 (sd = 6.82) and 31,282 (63.23%) males with a mean ventilation time of 4.41 (sd = 6.41). There was a significant difference between the two sexes (*χ*^2^ *p*-value < 2*e* − 16). When investigating age, 4,355 patients (8.80%) aged 18 to 39 had a mean ventilation time of 4.91 (sd = 6.47), 38,009 (76.84%) patients aged 40 to 79 had a mean ventilation time of 4.54 (sd = 6.61), and 7,103 patients aged 80 and above had a mean ventilation time of 4.51 (sd = 6.36). There was a significant difference found between the age groups (*χ*^2^ *p*-value = 3*e* − 08). The admission sources with the highest ventilation times included Other 373 (0.75%), with a mean ventilation time of 7.20 (sd = 8.52), and downstream 2,971 (6.00%), with a mean ventilation time of 6.93 (sd = 8.16). There was a significant difference found between the sources of admission (*χ*^2^ *p*-value < 2*e* − 16). There was also a significant difference found between the diagnosis, referring physician, and patient category groups with a *χ*^2^ *p*-value < 2*e* − 16. Patients with trauma (1,927 (3.90%)) had a mean ventilation time of 6.34 (sd = 7.13)) which was the longest average ventilation time and Cardiovascular patients (22269 (45.02%) had a mean ventilation time of 2.87 (sd = 4.77), which was the shortest. The MODS and NEMS scores were both significantly associated with ventilation time (*χ*^2^ *p*-value = 3*e* − 08). Pre-ICU hospital stay was significantly associated with ventilation time (*χ*^2^ *p*-value < 2*e* − 16). The MODS and NEMS scores were both significantly associated with ventilation time (*χ*^2^ *p*-value < 2*e* − 16). Scheduled admission and surgery were both significantly associated with ventilation time (*χ*^2^ *p*-value < 2*e* − 16). Among the recorded treatments received at arrival, only basic monitoring had no significant association with ventilation time (*χ*^2^ *p*-value =0.8). This can be explained by the fact that most of the patients (99.93%) received basic monitoring. This covariate was removed in further analysis.

In Figures 3.2 and 3.2, the log-rank test of the difference shown in Table 7 is confirmed with the distinction in the ventilation time of the various categories for each covariate.

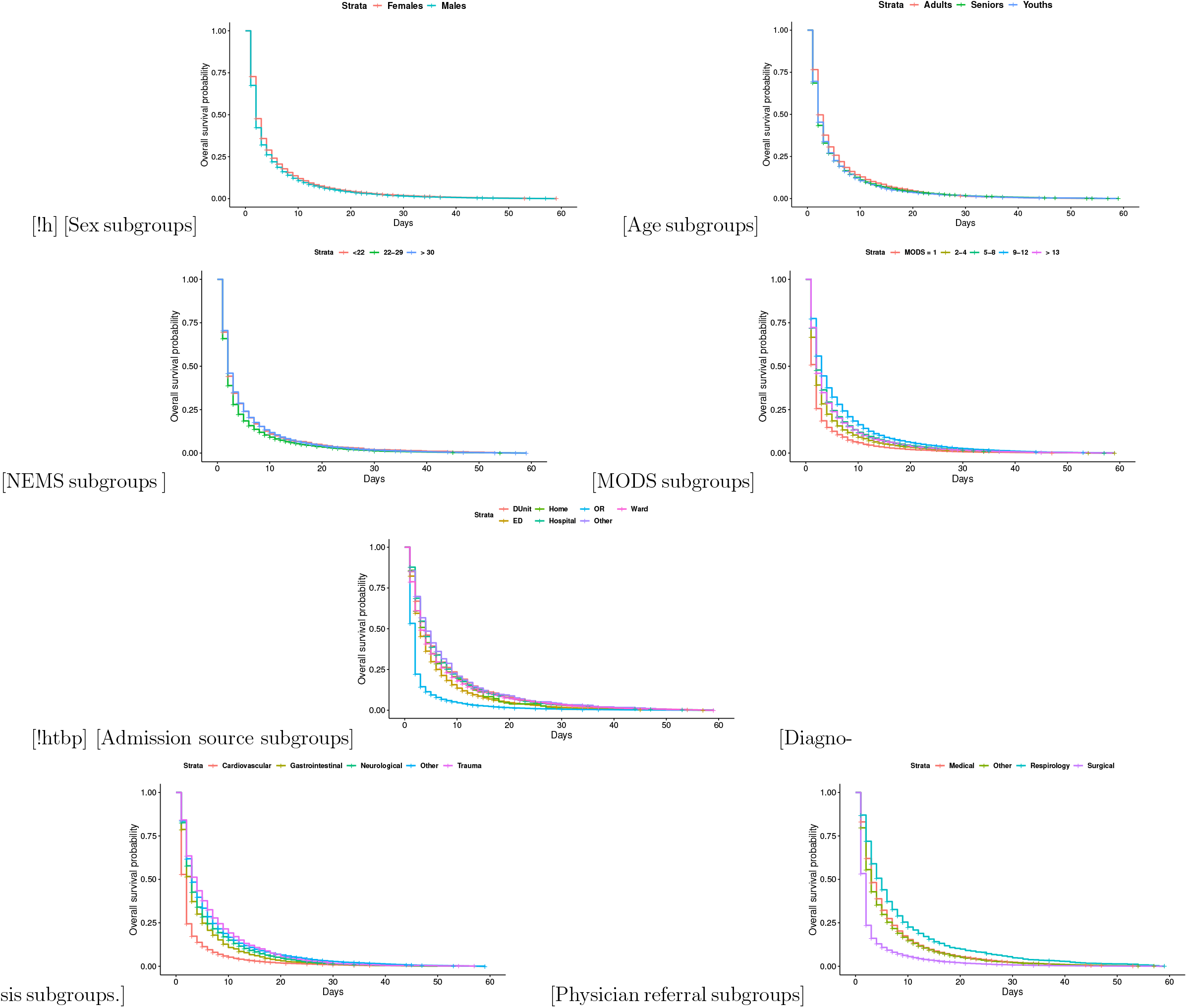

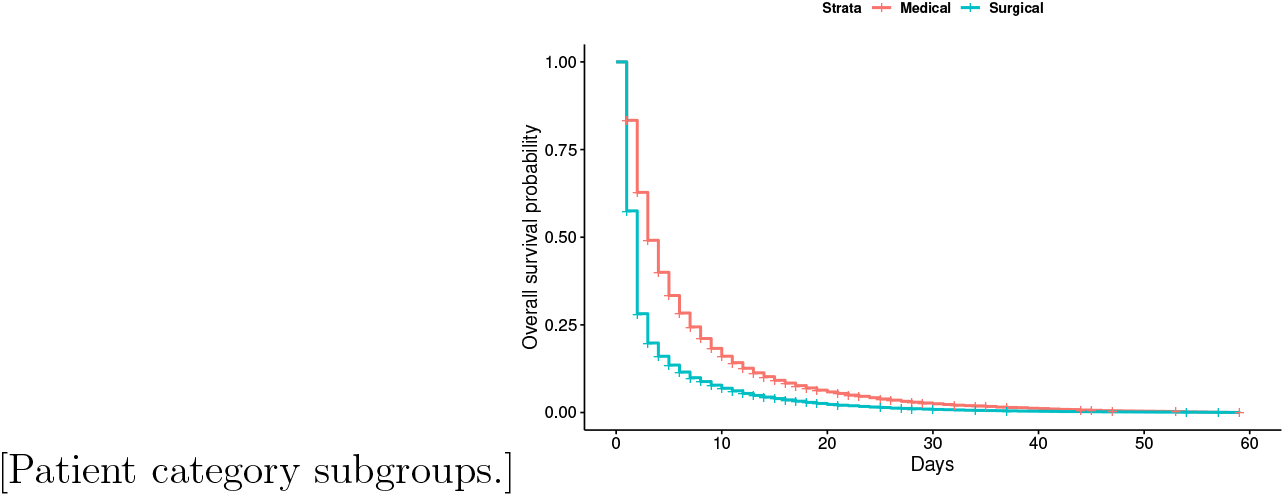

### 3.3 Probabilistic Characterization of ICU Ventilation Time

This study included a parametric analysis of ventilation time to determine the distribution with the best fit to the data. Figure 3.3(a) shows a plot of the negative log of the estimated survivor function against time. It shows approximately linear curve trends with minor deviation at the extremes. This suggests that the exponential distribution might be a good candidate. Figure 3.3(b) shows the plot of the log of the negative log of the estimated survivor function against log time. It shows an approximately linear trend and suggests that the Weibull distribution should be considered a good candidate. Figure 3.3(c) shows the plot of the cumulative probabilities versus log time. A concave trend can be observed with a faulty linear fit, suggesting that the log-normal distribution should be investigated further. Figure 3.3(d) shows the log of the survival probability versus the log of time in black, with a fitted linear model. It shows that the linear trend does not fit appropriately. This implies that the logistic model is not a good fit for the ventilation data.

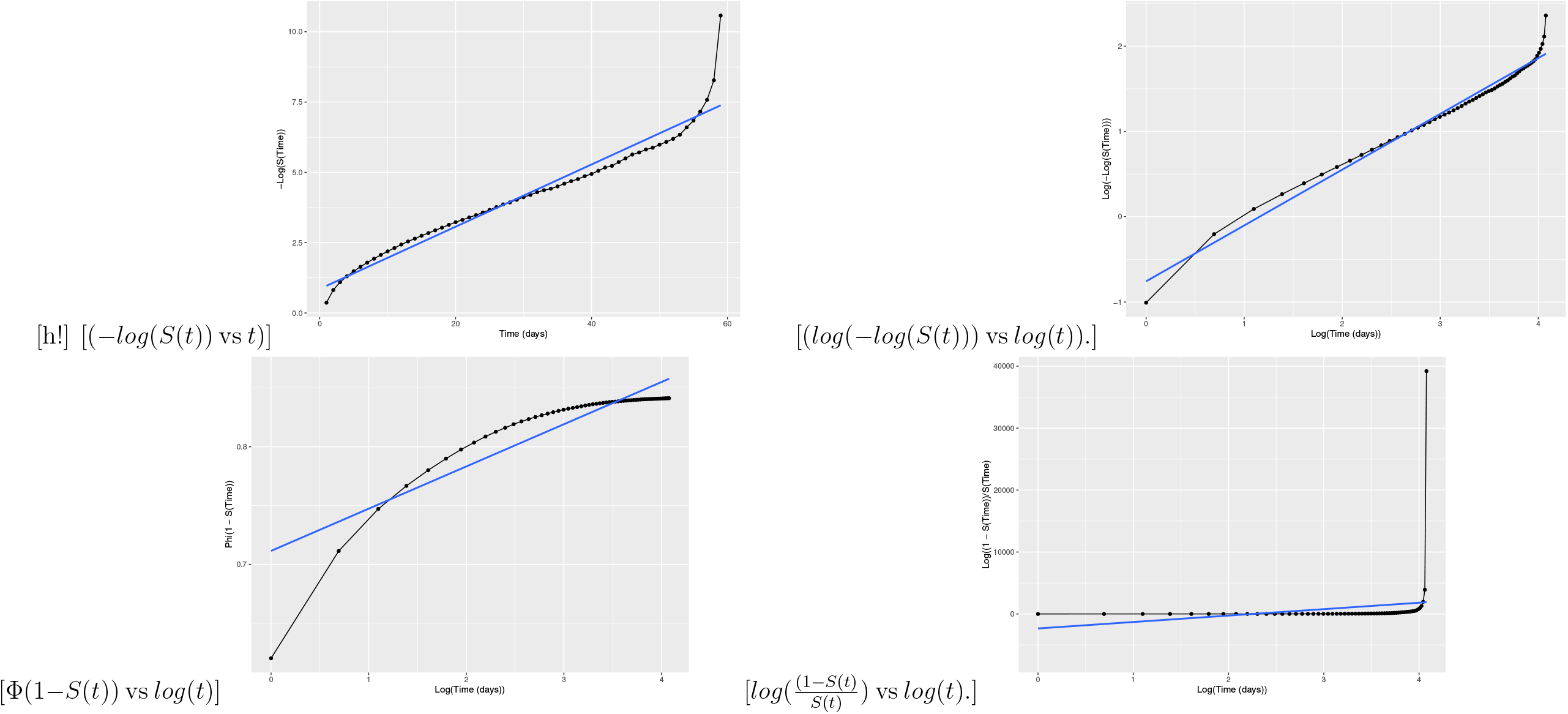

Figure 3.3(a) shows a histogram of the data overlaid with the density plot of the fitted distribution and Figure 3.3(b) contains the PP plot. Table 8 tabulates the criteria (Kolmogorov-Smirnov score (K-S), Cramer-von Mises score (C-M)), Anderson-Darling score (A-D), log-likelihood (log-l), the AIC, and the BIC) of the fitted distributions. Based on the criteria, the log-normal probability distribution function was identified as the best distribution for the First-day ventilation time. The maximum likelihood estimates of the shape and scale parameters and their standard deviations are given as 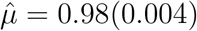, and 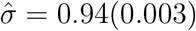 respectively. The actual form of the probability density function is:

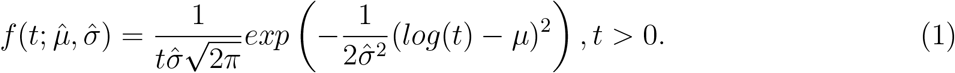

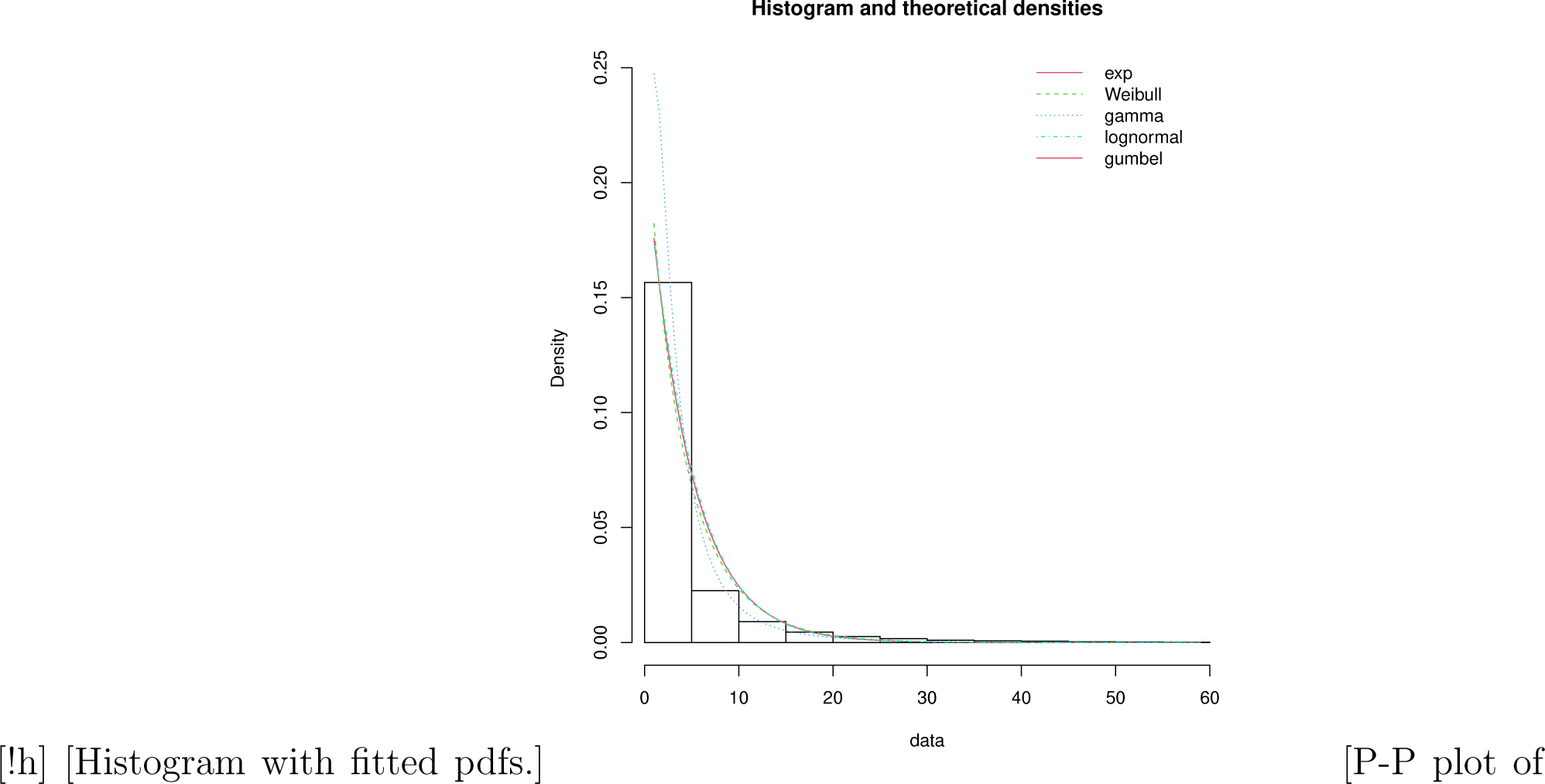

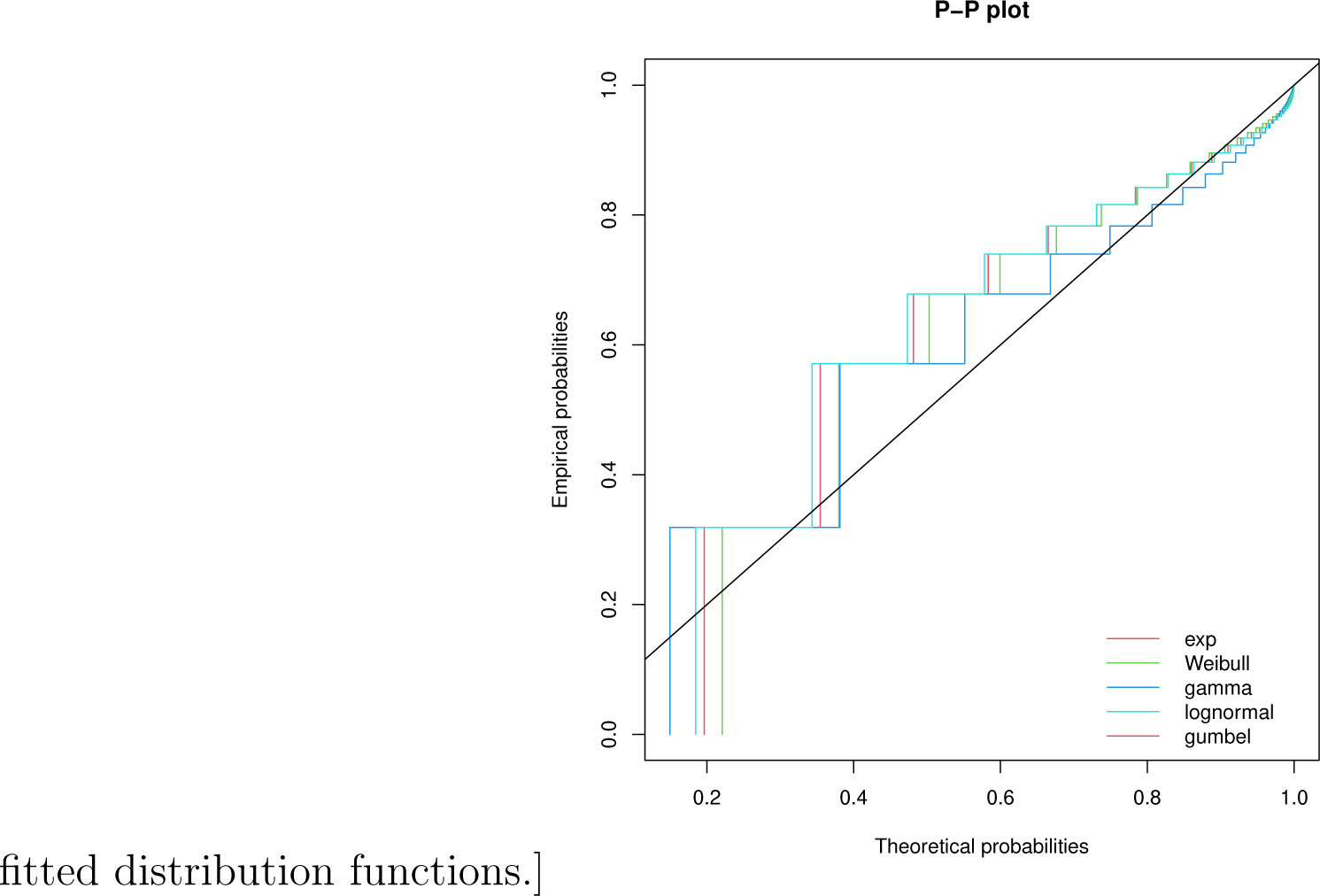

**Table 8:** MLE Estimates of ventilation time of all First-day ventilated patients.

| Dist | Estimate (sd) | K-S | C-M | A-D | log-l | AIC | BIC |
| --- | --- | --- | --- | --- | --- | --- | --- |
| Exp | $\hat{\lambda} = 0.22$ (0.00098) | 0.22 | 521 | 2929 | -124637 | 249275.3 | 249284 |
| Weibull | $\hat{k} = 0.94$ 0.003, $\hat{\lambda} = 4.40$ (0.02) | 0.22 | 462 | 2710 | -124409 | 248822 | 248840 |
| Gamma | $\hat{\sigma} = 1.06$ (0.01), $\hat{\lambda} = 0.23$ (0.002) | 0.23 | 555 | 3063 | -124588 | 249179 | 249197 |
| **Lognorm** | **$\hat{\mu} = 0.98$ (0.004), $\hat{\sigma} = 0.94$ (0.003)** | **0.19** | **299** | **1917** | **-115538** | **231080** | **231098** |
| Gumbel | $\hat{\alpha} = 2.46$ (0.01), $\hat{\sigma} = 2.78$ (0.01) | 0.26 | 739 | 4292 | -137551 | 275107 | 275124 |

Dist (distribution), sd (standard deviation, KS (Kolmogorov-Smirnov), C-M (Cramer-von Mises), and A-D (Anderson-Darling)

### 3.4 Cox Proportional Hazard Model

This section describes the fitting of the Cox-proportional hazard (PH) model to check the proportional hazard assumption. The Cox PH model makes no assumption about the distribution of the event time, but it assumes that the hazard ratio is constant over time. This assumption was tested for each covariate and globally. The results of the goodness of fit test of the proportional hazard assumption, which are tabulated in Table 15 in the Appendix, give a significant *p*-value < 2*e* − 16. Therefore the null hypothesis that the proportionality assumption holds is rejected globally, indicating a lack of proportionality for the hazard function. There is a significant deviation from the proportional hazard assumption for all the variables (*p*-value < 0.05). By inspecting Figure s 3.2 and 3.2, the lines for male and female patients as well as the various age groups were not parallel, confirming that the proportional hazard assumption is not reasonable in this case involving of stratified data. The PH model is not appropriate for these data and the ventilation time was therefore modelled using the AFT model.

### 3.5 Accelerated Failure Time Model

#### 3.5.1 Model Selection

To model the ventilation time, Exponential, Weibull, Log-logistic, and Lognormal AFT models were fitted to the data. In each case, the model was fitted to all the covariates without Basic monitoring on arrival. From Table 9, each model was assessed using the Akaike Information Criterion (AIC), the Bayesian Information Criterion (BIC), and the Log-Likelihood model from the selection process. The Log-logistic model (log-likelihood = -73,400.5, AIC = 147,067, and BIC = 148,191) was a better fit, as it is the model with the smallest criteria. The fitted log-logistic AFT model on all covariates is tabulated in Table 16 in the Appendix.

**Table 9:**
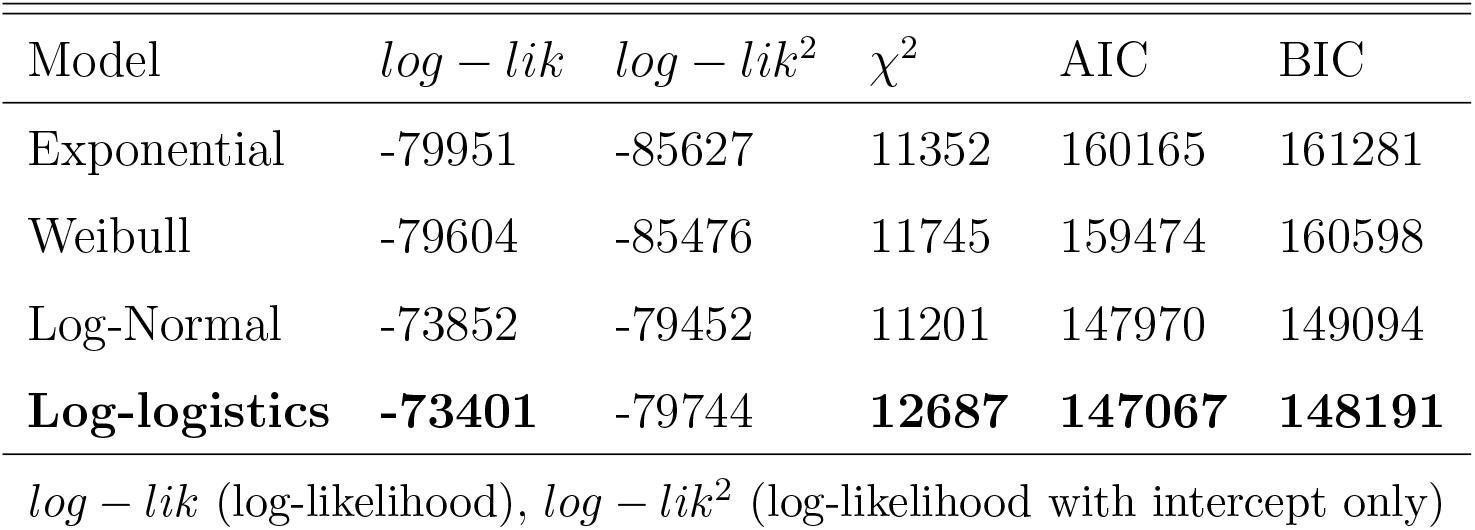
Performance comparison of AFT models on the regional data

The goodness of fit of each model was assessed using the distribution of the Cox-Snell residuals. To do this, we compared the survival estimates of each parametric model were compared with the KM estimates, by plotting the survival probability against the Cox-Snell residuals. The survival function should closely follow the KM estimate. From Figure 3.5.1, the survival function for the Log-logistic model in Figure 3.5.1(d) is superimposed on the KM curve, clearly showing that this model approximates the empirical survival better than the other models.

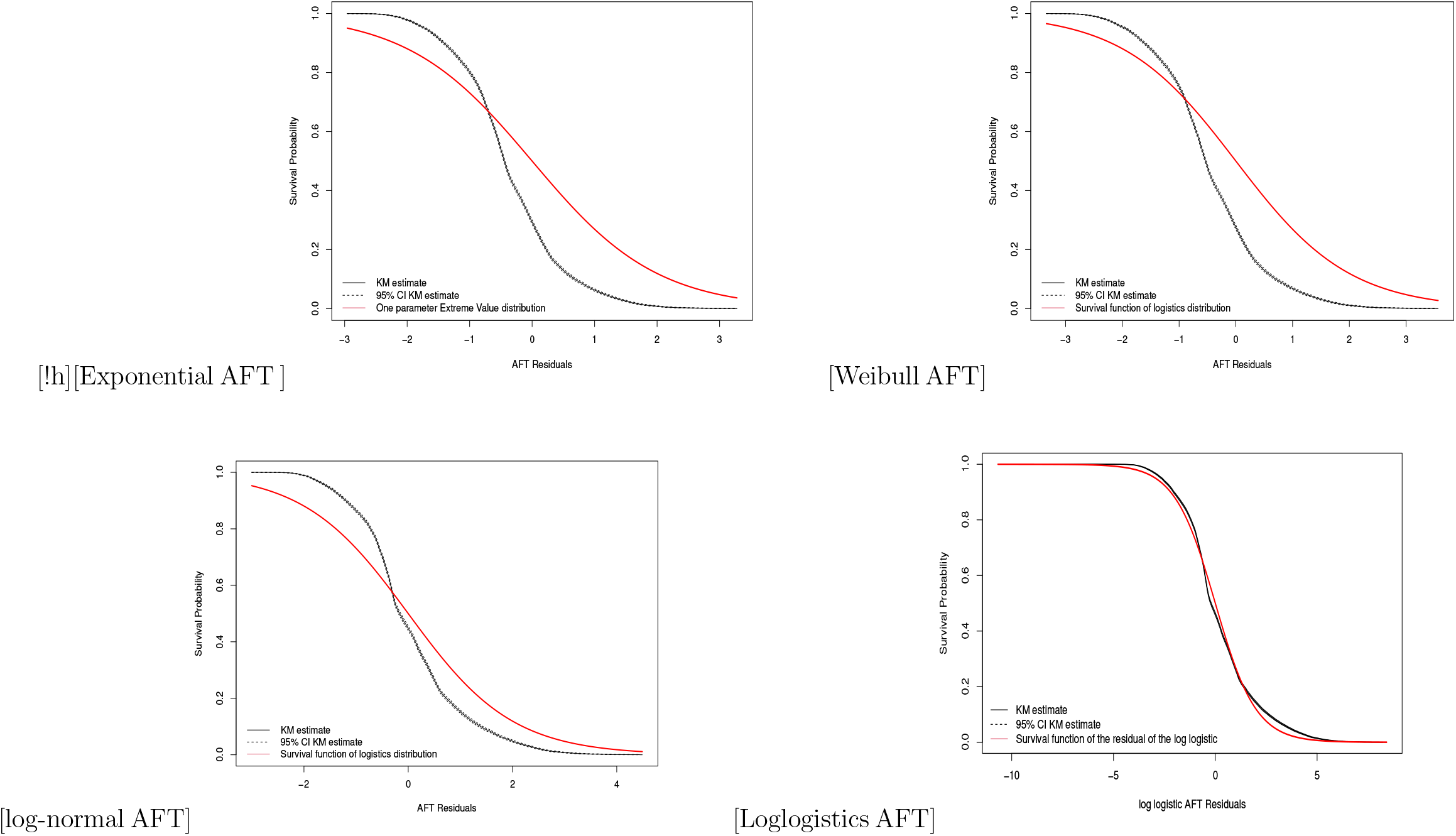

#### 3.5.2 Variable Selection

Variable selection followed the approach outlined in the methods section. Using the backward selection procedure, patient category (*p*-value > 0.74), dialysis (*p*-value > 0.57), interventions outside (*p*-value > 0.10) and gender (*p*-value > 0.65) were eliminated, resulting in the final model presented in Table 10.

**Table 10:**
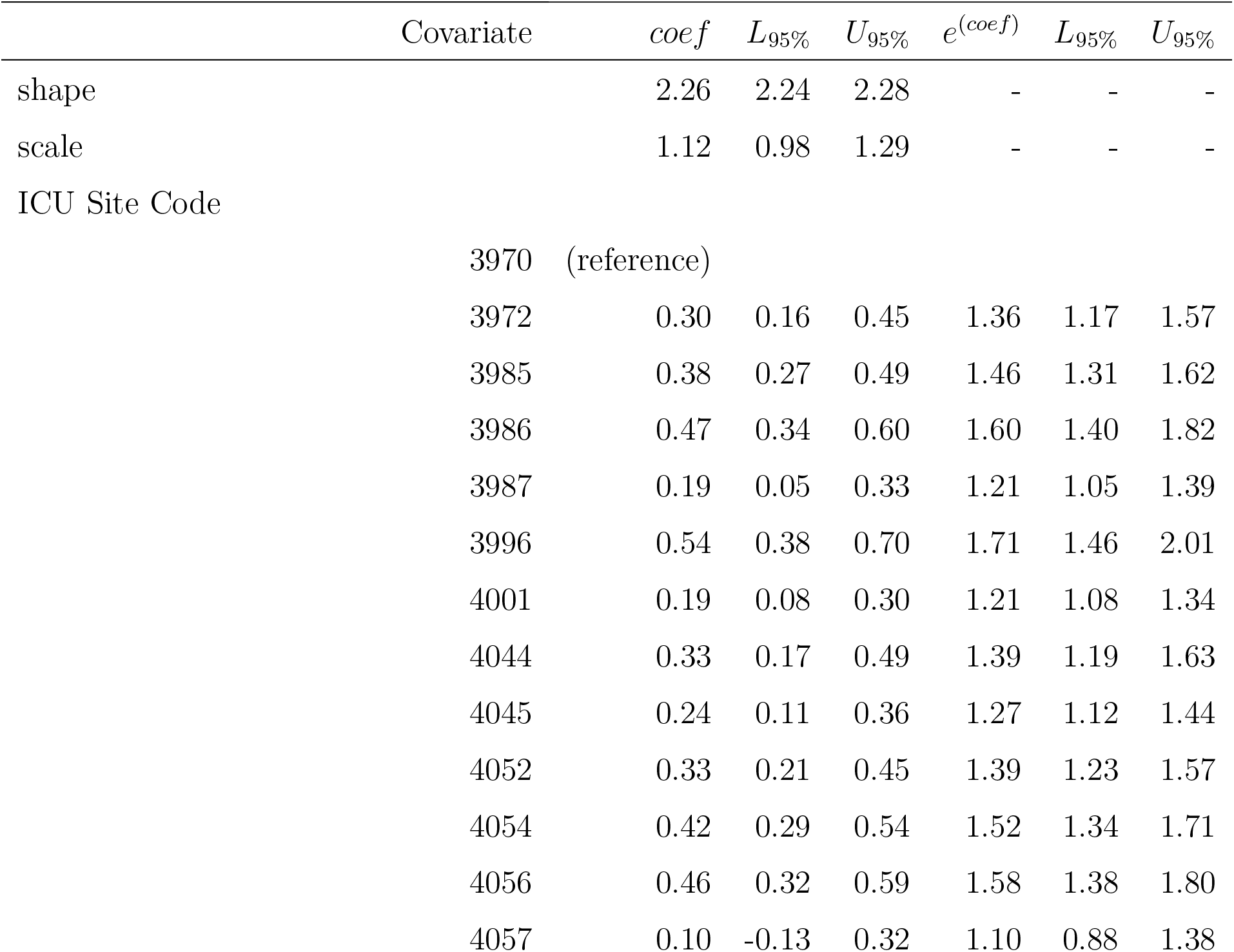
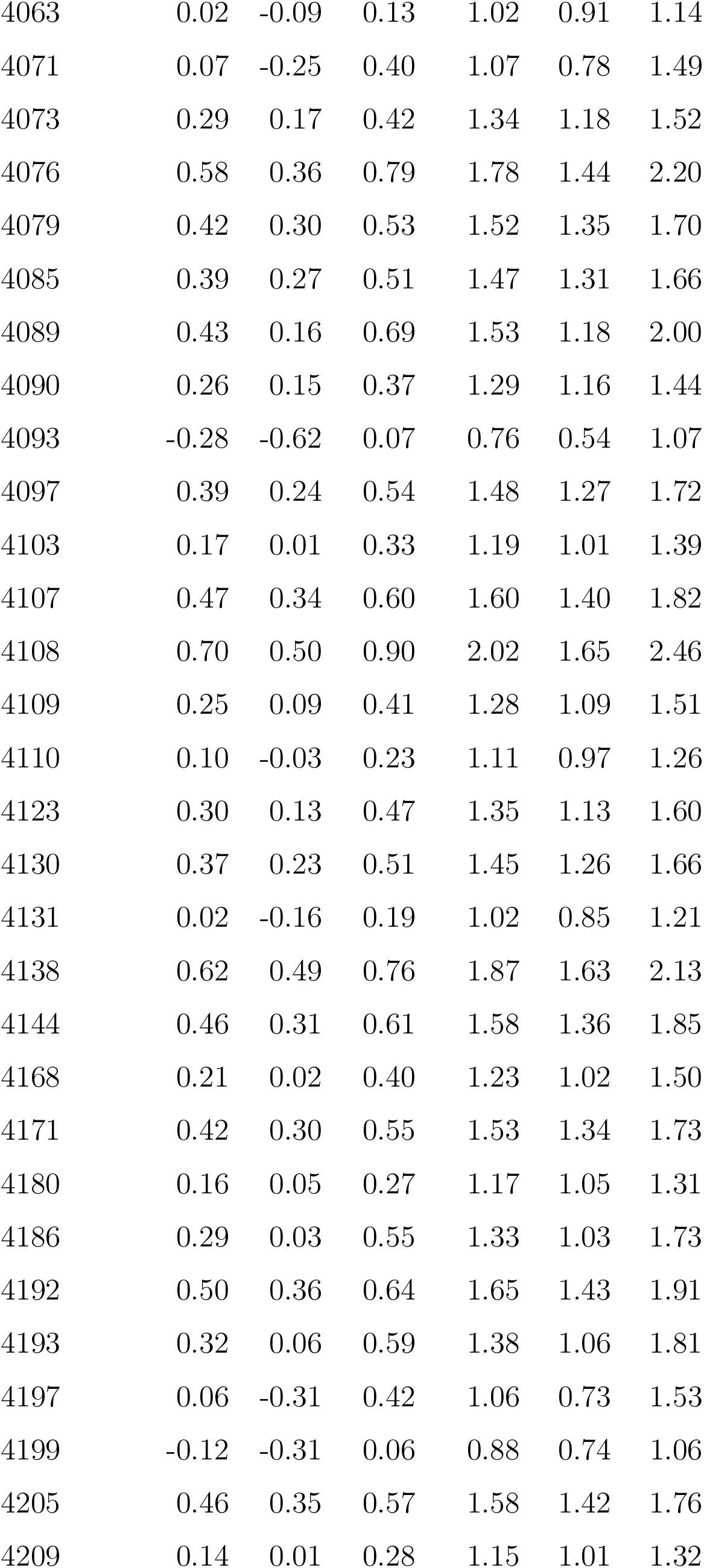
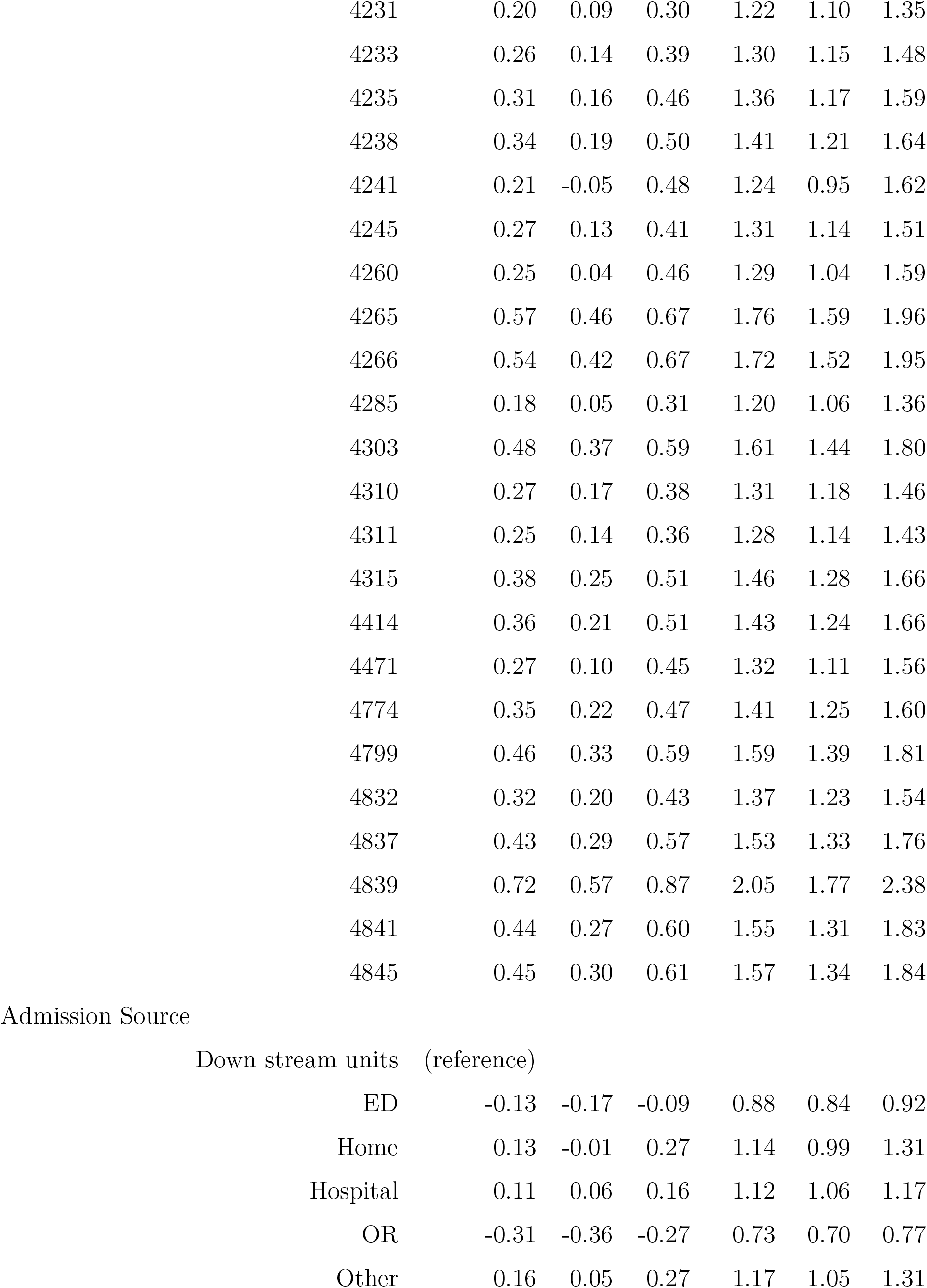
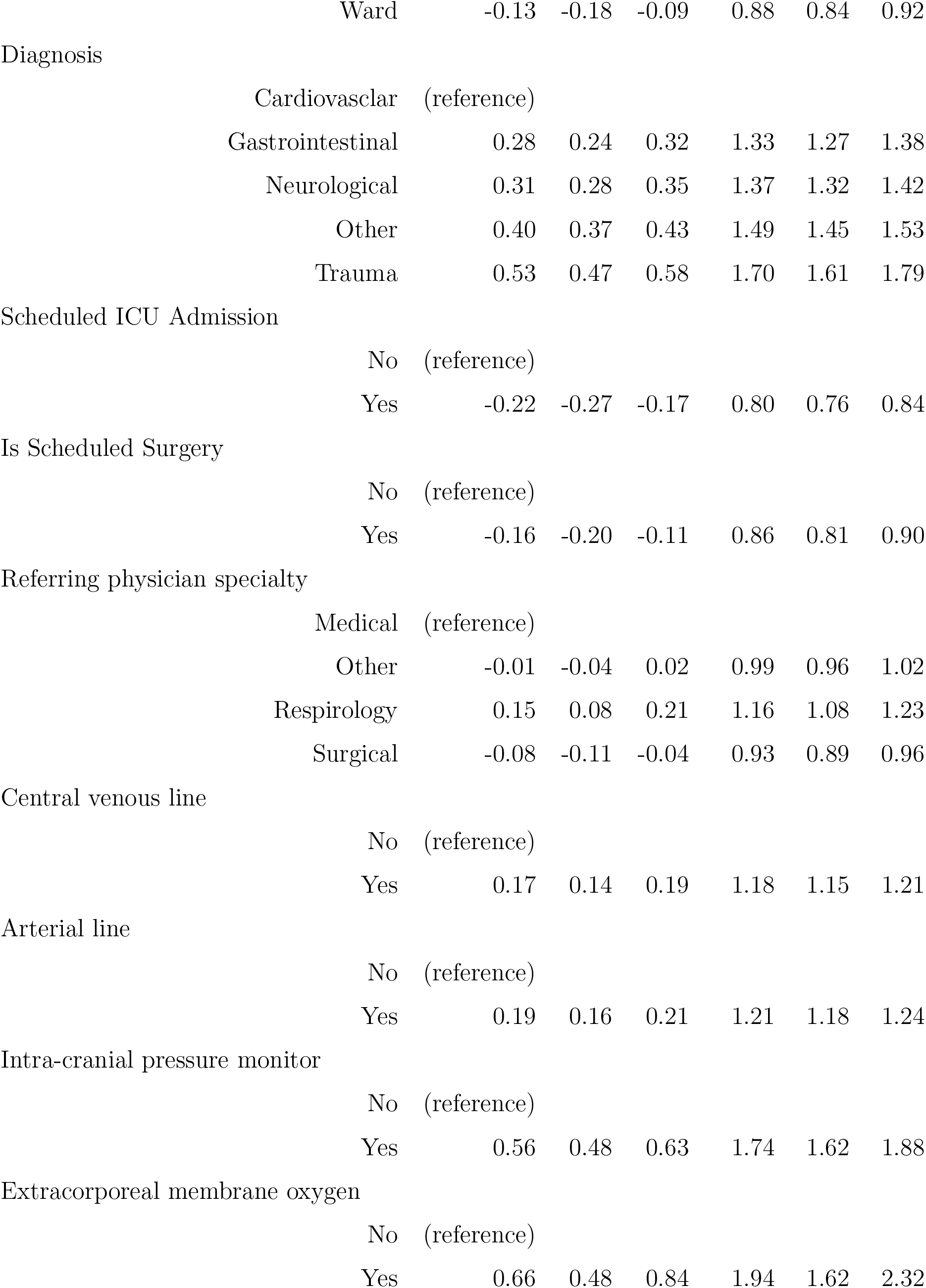
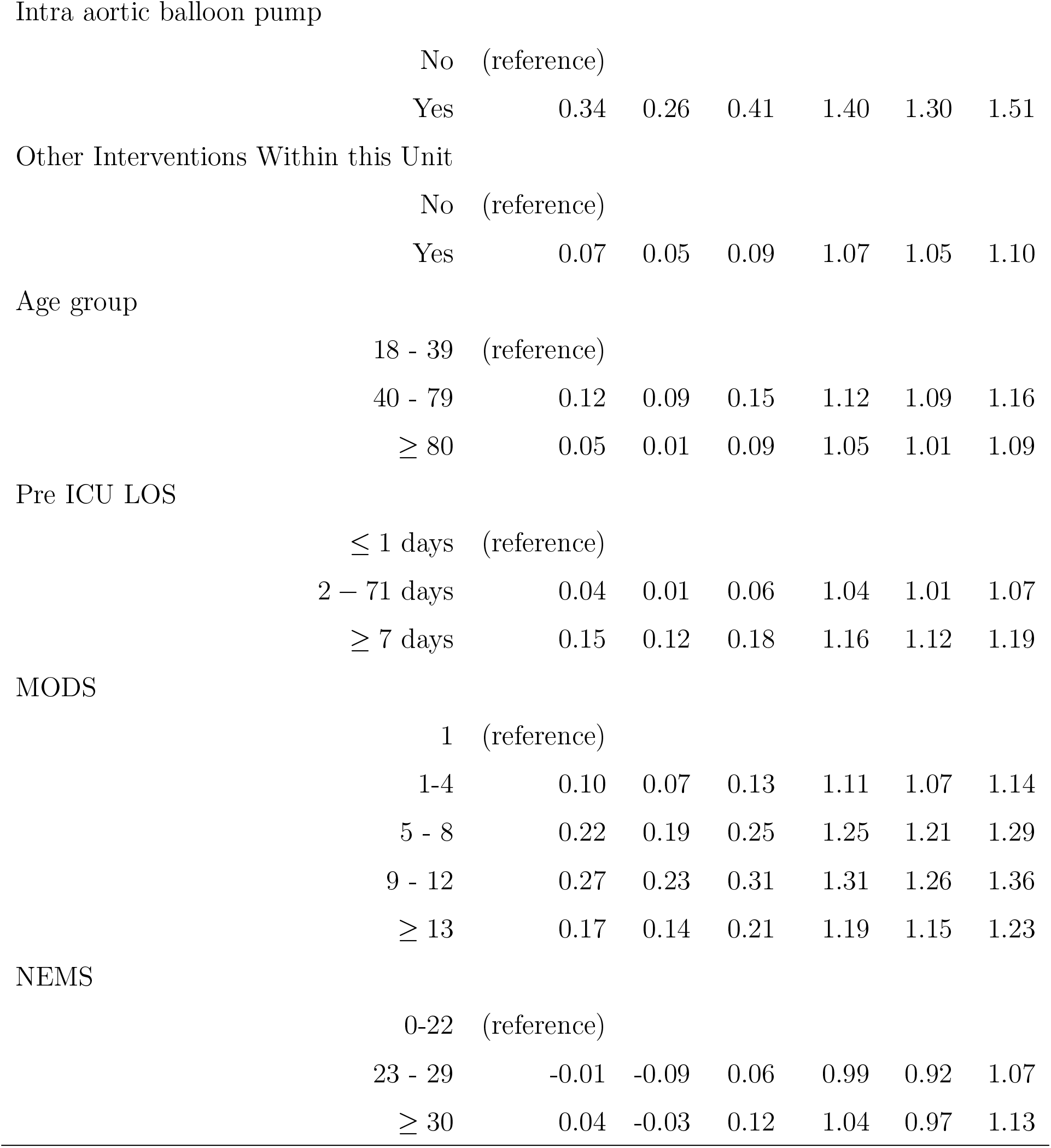
Log-logistic AFT model of the training set

Analysis with the log-logistic AFT model revealed that the model containing the explanatory variables significantly improved the predictive ability of the model with the intercept only, because the likelihood ratio gave a *p*-value < 2*e* − 16. The overall effect of each of the retained covariates on survival time revealed that all had a significant independent effect on IMV time (all likelihood ratio tests resulted in a *p*-value < 10*e* − 5). Table 17 tabulates the likelihood ratio test results of variable selection criteria for models fitted to the data using backward elimination.

#### 3.5.3 Model predictive performance

To assess the predictive performance of the model, a test dataset was used to predict the ventilation time using the log-logistic model. The residuals of the test data were computed and compared with those of the training data. Table 11 presents a comparison of predictive performance on the training and testing data. The predicted average ventilation duration was 2.96 days for the test data compared with 2.94 days for the training data. There were insignificant losses of performance from the prediction of the training data to that of the test data in the quantiles (1.60, 2.75, 3.81 days to 1.59, 2.77, 3.85 days), in the MSE (from 46.81 to 60.21), in the MAE (from 2.95 to 3.00), in the PBIAS (from 0.36 to 0.35), and in the NES (from 1.090 to 1.380).

**Table 11:** Comparison of prediction statistics from the test and training datasets.

| Statistics | N (%) | Mean | SD | $Q_{0.25}$ | $Q_{0.5}$ | $Q_{0.75}$ | MSE | MAE | PBIAS | NES |
| --- | --- | --- | --- | --- | --- | --- | --- | --- | --- | --- |
| Training | 34,626 (70%) | 2.94 | 2.83 | 1.59 | 2.77 | 3.85 | 46.81 | 2.95 | 0.36 | 1.09 |
| Test | 14,840 (30%) | 2.96 | 4.55 | 1.60 | 2.75 | 3.81 | 60.21 | 3.00 | 0.35 | 1.38 |

Figure 3.5.3 shows the survival functions of the training (in blue) and testing (in black) residuals superimposed on the baseline KM survival. The survival curves of the training and testing data are similar to the baseline KM survival. The performance of the training model is revealed on the test data with an insignificant difference and a narrow gap. This indicates that the model has a good ability to predict patients’ ventilation duration using First-day observations.

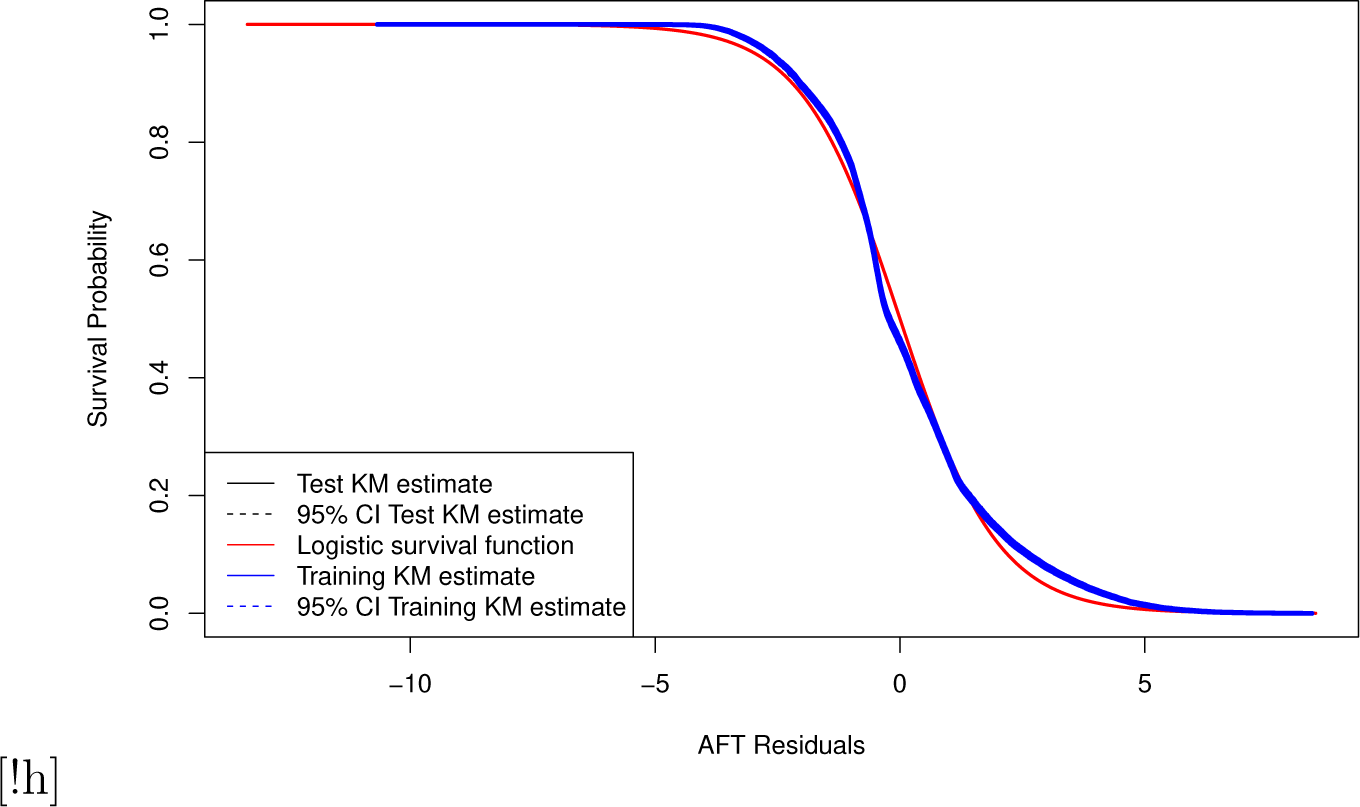

#### 3.5.4 Model Validation

To validate the model predictions, a new dataset from the London Health Sciences Center (LHSC), with data gathered from January 2019 to May 2021 was used. The first step was to calibrate the data. Calibration checks the agreement between observed and predicted outcomes. To assess the need for model calibration, simple linear regression and scatter plot of observed versus predicted outcomes was performed. Perfect predictions would yield a slope of 1 with an intercept of 0, which is the line of best fit that should divide the first quadrant into two equal parts. A failure could inform the need for a model that considers shrinkage. The predictive performance compares the estimated Kaplan-Meier survival curve of the predicted residual to the expected empirical survival curve. This was done on three subgroups: COVID-19 patients, Non-COVID-19 patients, and the whole LHSC dataset. Figure 3.5.4(a) shows the residual of the survival obtained from the LHSC data from January 2020 to May 2021, including all patients that received IMV on arrival.

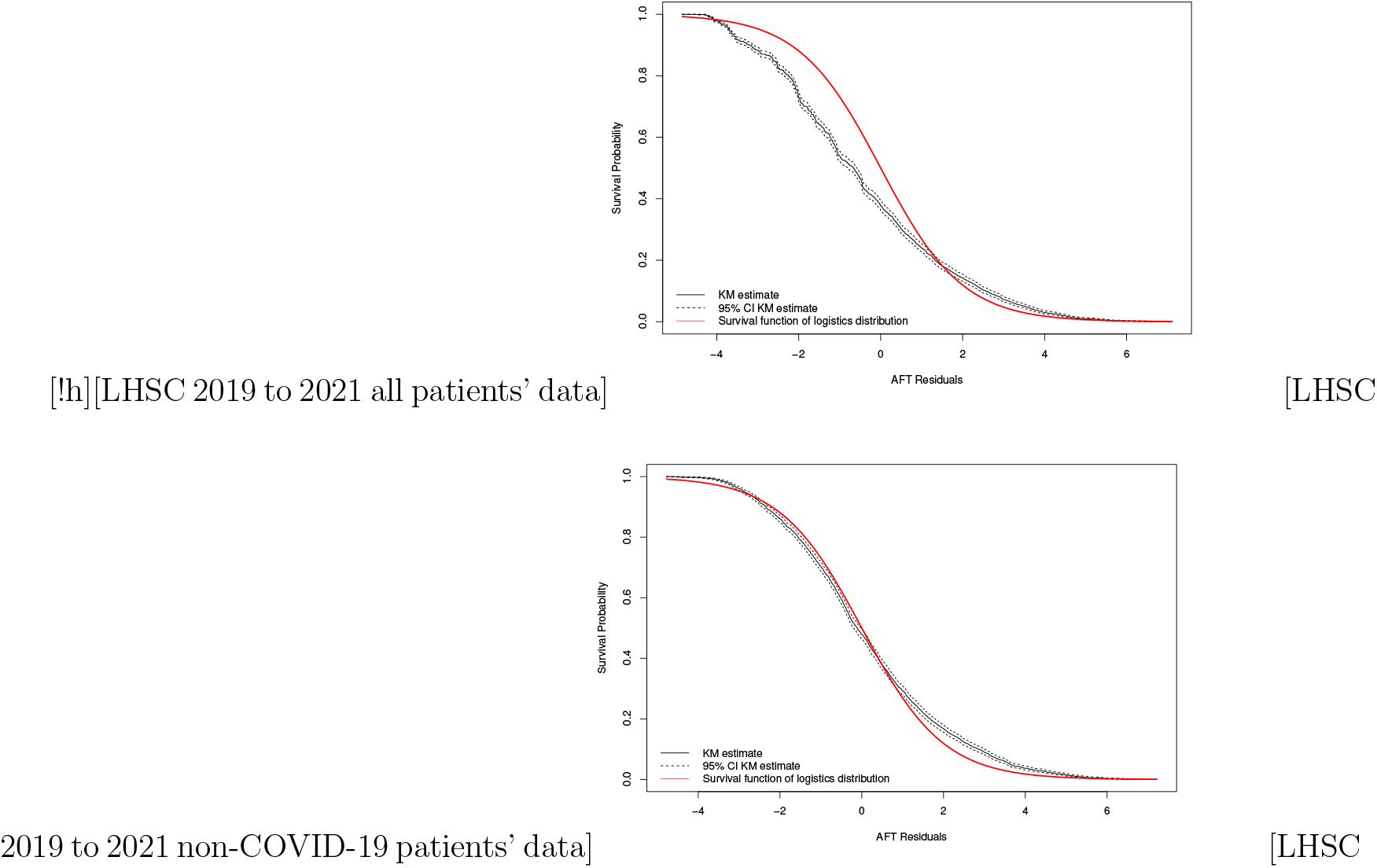

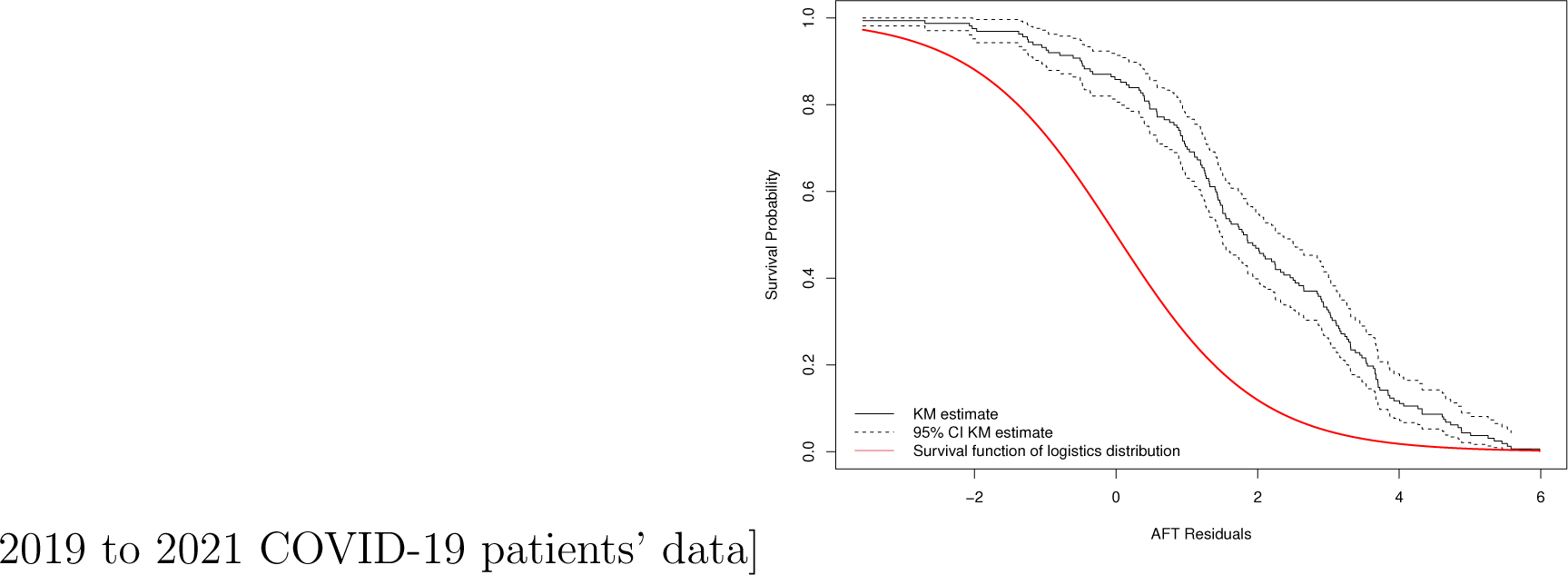

In Figure 3.5.4(a), the survival curve of the AFT model poorly approximates the expected empirical survival, there appears to be a major departure between the two. This could have been due to a medical condition that was not present in the previous time interval, in particular, COVID-19. To confirm this assumption about the effect of COVID, the data were divided into two subsets: patients with COVID-19, and patients without COVID-19.

Interestingly, in Figure 3.5.4(b), the survival function closely follows the KM estimate, suggesting a good fit for the non COVID patients. However, the survival plot of the residual from the prediction of the COVID patients in Figure 3.5.4(c) performed poorly. This is confirmed in Table 12, where the model’s predictive performance on the three sub-datasets (Covid patients, Non-COVID patients, and Mixed data) is numerically compared.

**Table 12:**
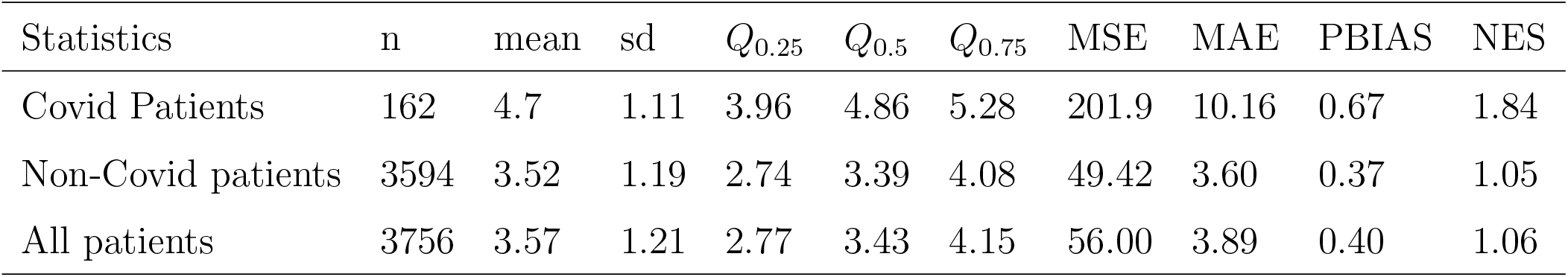
Model validation performance on three sub-data set.

| Statistics | n | mean | sd | $Q_{0.25}$ | $Q_{0.5}$ | $Q_{0.75}$ | MSE | MAE | PBIAS | NES |
| --- | --- | --- | --- | --- | --- | --- | --- | --- | --- | --- |
| Covid Patients | 162 | 4.7 | 1.11 | 3.96 | 4.86 | 5.28 | 201.9 | 10.16 | 0.67 | 1.84 |
| Non-Covid patients | 3594 | 3.52 | 1.19 | 2.74 | 3.39 | 4.08 | 49.42 | 3.60 | 0.37 | 1.05 |
| All patients | 3756 | 3.57 | 1.21 | 2.77 | 3.43 | 4.15 | 56.00 | 3.89 | 0.40 | 1.06 |

## Discussion

Accurate prediction of ICU resources helps guide therapeutic decision-making, resource allocation, and patient flow management. The number of days on IMV is a major concern of critical care management and costs [Dasta et al., 2005, Bice and Carson, 2019]. However, IMV duration prediction models in the literature were mainly based on the conventional multivariate regression model and logistics regression but did not incorporate censored observations and were based on classifying patients’ ventilation time as either short or long duration [Troche and Moine, 1997, Dimopoulou et al., 2003, Abujaber et al., 2020, Figueroa-Casas et al., 2015, Trouillet et al., 2009, Aung et al., 2018]. A comprehensive survival analysis was performed to predict and determine predictors of ventilation time using the CCIS Ontario dataset. Information obtained at arrival is an important component of forecasting patient ventilator days.

The Log rank test on the KM curves of the covariates available on the first-day of ventilation show that only basic monitoring was insignificant (p-value = 0.80). This is likely because only a very small number did not receive the basic monitoring treatment and therefore do not have the power to rule out a real difference and avoid a type two error (false negative).

The covariate’s effect in the AFT model is to accelerate or decelerate the event time, which in this case is the invasive ventilation time. The results of the association are shown in Table 10. A convenient way to understand the coefficients better is through the interpretation of the time ratio (TR), also called the acceleration factor. The TR for a given covariate is the (natural) exponent of the estimated parameter coefficient (i.e, *exp*(*β*)). A positive coefficient corresponds to a TR greater than 1, whereas a negative coefficient corresponds to a time ratio less than 1. Correspondingly, a TR greater than one implies that the covariate increases the time to the event. An acceleration factor equal to 1 corresponds to no effect on the time to the event.

A positive coefficient was observed for most of the ICUs (SiteCode), implying that for most ICUs, the time to event was higher than average. In relative terms, the ICU that a patient visited was a significant predictor of the patient’s ventilation time. Different ICUs had differing TR. Compared to the ICU with code 3970 (used as a reference), ICUs with site codes 4057, 4063, 4071, 4093, 4110, 4131, 4197, 4199, and 4241 (9 out of 65) had an insignificant coefficient. In other words, their acceleration factor’s confidence interval covers 1 and is not significantly different from that of 3970. Moreover, the 56 remaining ICUs had significantly higher TR. Attending one of those ICUs increased the time that a patient spent using a ventilator. This large difference in ventilation time in the ICU may be attributed to the practices and locations of the various ICUs as outlined in Burns et al. [2021]. The implication of this for ICU management is that ICU managers should learn best practices from the ICUs that seem to have a lower ventilation time.

Comparing patients admitted to the ICU from the downward stream (SDU and Level 3) to those patients admitted from the house (TR = 1.14, CI = (0.99,1.31)), there was no significant difference. However, patients from the ED (TR = 0.88, CI = (0.84 0.92)), OR (TR = 0.73, CI = (0.70, 0.77)), and ward (TR = 0.88, CI =(0.84, 0.92)) had lower odds of longer ventilation time, whereas patients admitted from hospitals (TR = 1.12, CI = (1.06, 1.17)) and other sources (TR = 1.17, CI = (1.05,1.31)) had higher odds of longer ventilation time within the ICU. Those admitted with cardiovascular, cardiac, vascular diagnoses had experienced longer ventilation time as compared to those with other etiologies. Patients with gastrointestinal (TR = 1.33, CI = (1.27 1.38)), Neurological (TR = 1.37, CI = (1.32 1.42)), Trauma (TR = 1.70, CI = (1.61 1.79)), and Other diagnoses (TR = 1.49, CI = (1.45 1.53)) had higher odds of staying on the ventilator longer than cardiovascular patients. This could be explained by the findings of Kao et al. [2016] using the same data where patients with cardiovascular diagnoses had higher mortality than those with other etiologies (Priestap et al. [2020]). Patients’ admission diagnosis types were also significant factors affecting ventilation time. Surgical patients (TR = 0.770, CI = (0.702, 0.843)) had higher odds of leaving the ventilator earlier than medical patients.

Scheduled ICU admission (TR = 0.827, CI = (0.784, 0.873)) had a shortening effect on ventilation time. This implies that scheduled admission is a significant predictor of patients’ ventilation time. Scheduled patients have 17.3% higher odds for shorter ventilation time compared to non-scheduled patients. This may be attributed to the fact that generally, scheduled patients are taking medication or receiving treatment that supports their fast and safe transit through the ventilation, and are therefore less likely to remain connected to the ventilator for long periods than non-scheduled patients. Moreover, compared to non-scheduled surgery patients, patients with scheduled surgery (TR = 0.823, CI = (0.783, 0.864)) had a 17.7% higher probability of shorter ventilation time. Referring Physician services such as Cardiology (TR = 1.100, CI = (0.989, 1.220)), Ophthalmology (AF = 1.39, CI = (0.726, 2.64)), and Psychiatry (AF = 1.01, CI = (0.639,1.610)) had non-significant effects and acted as an average baseline. However, the rest of the referring physicians’ services had a significantly higher TR, with none that were significantly lower. For example, patients from Trauma (TR = 2.28, CI = (2.12, 2.45)) and Paediatric (TR = 10.100, CI = (2,910, 34.900)) had the longest time on a ventilator.

Unequivocally, treatments received on arrival had a significant effect on ventilation time. The likelihood of longer ventilation was much higher for patients who received a central venous line (CVL) (TR = 1.18, CI = (1.15, 1.21)), arterial line (AL) (TR = 1.21, CI = (1.18, 1.24)), intracranial pressure monitor (IPM) (TR = 1.74, CI =(1.62, 1.88)), extracorporeal membrane oxygen (EMO) (TR = 1.94, CI = (1.62, 2.32)), Intra Aortic Balloon Pump (IABP) (TR = 1.40, CI = (1.30, 1.51)), and other intervention within the ICU (OIWU) (TR = 1.07, CI =(1.05, 1.10)). These treatments acted to decelerate the event. Patients who received these interventions in the ICU on arrival were connected for a longer time than compared to those who did not. These treatments are significant for predicting a patient’s ventilation time.

The odds of longer ventilation time are much higher for adults and seniors than for younger people. Patients aged 40–79 (TR = 1.12, CI =(1.09, 1.16)) and those *≥* 80 years old (TR = 1.05, CI = (1.01, 1.09)) spent longer time on the ventilator than patients aged *≤* 39. This confirms the results reported by Piotto et al. [2012] and Lei et al. [2009] who showed that advanced age (more than 60 years) was a significant predictor for IMV. However, this study found that the patient’s sex was not a significant predictor. Longer pre-ICU LOS was associated with longer ventilation time. Specifically, patients who spent more than one day but less than seven in the hospital post-ICU (TR = 1.04, CI = (1.01, 1.07)), and those who spend more than a week post-ICU admission (TR = 1.16, CI =(1.12, 1.19)) are more likely to experience the event later than those who spent less than one-day post ICU admission. Higher First-day scores in MODS corresponded to increasing time to event (MODS = (1-4), TR = 1.11, CI = (1.07 1.14)) (MODS = (5 - 8), TR = 1.25, CI = (1.21 1.29)), (MODS = (9 - 13), TR = 1.31, CI = (1.26 1.36)), and (MODS *≥* 13, TR = 1.19, CI = (1.15 1.23)). Patients with high MODS scores upon arrival were at a higher risk of longer ventilation connection times than those with low scores.

he First-day NEMS score however had a weak association with ventilation time (NEMS = (0-22), TR = 0.99, CI = (0.92 1.07)), (NEMS *≥* 30, TR = 1.04, CI = (0.97 1.13)). In the review by Ghauri et al. [2019], results based on logistic regression models indicated no significant effects for the Acute Physiology and Chronic Health Evaluation (APACHE II) on the prediction IMV time of ICU patients. Although one would expect NEMS to be a significant predictor because of the weight of the respirator component, the results show the opposite, that it is not closely associated. However, the presence of the covariate in the model significantly increased the model’s predictive performance, as seen in the variable selection.

## Conclusion

This study proposed survival analysis to investigate the duration of invasive mechanical ventilation in ICU patients. Both parametric and non-parametric methods were explored. Based on the AIC and BIC criteria, the log-logistic AFT model was retained as the best predictive model of ventilation time for each ICU patient. ICU site, admission source, admission diagnosis, scheduled admission, scheduled surgery, referring physicians specialty, central venous line treatment, arterial line treatment, intracranial pressure monitor treatment, extra-corporeal membrane oxygen treatment, intra-aortic balloon pump treatment, other interventions, age group, pre ICU LOS, and MODS score were significant predictors of ICU ventilation time. Even though the data used for modelling were five years old, they performed well on current non-COVID patients. The predictive performance of the proposed model showed that it can be used to predict future ventilation time duration and provide insight into predictors of ventilation time.

This study differs from previous studies in several ways. First, unlike the studies of [Troche and Moine, 1997, Dimopoulou et al., 2003, Abujaber et al., 2020, Figueroa-Casas et al., 2015, Trouillet et al., 2009, Aung et al., 2018] this study focused entirely on predicting continuous ventilation time using information gathered on the first day. In addition, the sample size was larger and external validation was used instead of bootstraps. This translates into more power to detect predictive performance.

Nevertheless, this study has several limitations. The analysis included only patients who entered the ICU over one and one half years. Although the patient characteristics of this subgroup were not dissimilar to those of the patients in the validation set, it must be noted that the different study period could have significant, unrecognizable differences due to the appearance of COVID-19. The heterogeneity of the ICU sites may also have affected the results. Because the models performed differently for different ICUs, each ICU sites could be considered as a random effect. In that case, each ICU woull have to have its own model. Nonetheless, with the current model, the acceleration functions of the various ICUs in Ontario can be compared. The survival model used in this research could benefit from the automatic variable selection tools of penalized AFT models. In addition, other machine learning methodologies, such as the random forest and Support vector machines (SVM) could be used. In addition, the nonlinear tree based machine learning algorithms implemented in libraries such as XGBoost, scikit-learn, LightGBM, and CatBoost with higher accuracy estimation could be used. However, state-of-the-art implementations of such methods for the AFT models are few.

## Data Availability

Critical Care Information System datadase

## Acknowledgments

We would like to thank the LHSC staff for providing the data used in this analysis.

Author contributions:CPDS contributed statistical insight and edited the manuscript. FFR contributed health care managerial and clinical insight and edited the manuscript, and KYM worked on analysis, coding, drafting and writing of the paper. MW contributed to the literature review and the editing of the manuscript.

The R code used to conduct the analyses is available upon request without the data.

## Appendix

**Table 13:** Selected estimates from the K-M curve.

| Time | n.risk | n.event | n.censor | surv | UCI | LCI |
| --- | --- | --- | --- | --- | --- | --- |
| 1 | 49,467 | 15,125 | 646 | 0.694 | 0.698 | 0.690 |
| 2 | 33,696 | 12,213 | 273 | 0.443 | 0.447 | 0.438 |
| 3 | 21,210 | 5,179 | 129 | 0.335 | 0.339 | 0.330 |
| 4 | 15,902 | 2,979 | 60 | 0.272 | 0.276 | 0.268 |
| 5 | 12,863 | 2,092 | 39 | 0.228 | 0.231 | 0.224 |
| 6 | 10,732 | 1,594 | 38 | 0.194 | 0.197 | 0.190 |
| 7 | 9,100 | 1,270 | 28 | 0.167 | 0.170 | 0.163 |
| 8 | 7,802 | 998 | 33 | 0.145 | 0.149 | 0.142 |
| 9 | 6,771 | 866 | 26 | 0.127 | 0.130 | 0.124 |
| 10 | 5,879 | 698 | 20 | 0.112 | 0.115 | 0.109 |
| 15 | 3,249 | 326 | 8 | 0.064 | 0.066 | 0.062 |
| 20 | 1,963 | 176 | 0 | 0.040 | 0.041 | 0.038 |
| 25 | 1,258 | 110 | 6 | 0.026 | 0.027 | 0.024 |
| 30 | 791 | 68 | 2 | 0.016 | 0.018 | 0.015 |
| 35 | 528 | 41 | 2 | 0.011 | 0.012 | 0.010 |
| 40 | 334 | 26 | 0 | 0.007 | 0.008 | 0.006 |
| 45 | 201 | 25 | 1 | 0.004 | 0.005 | 0.004 |
| 50 | 119 | 12 | 0 | 0.003 | 0.003 | 0.002 |
| 55 | 56 | 12 | 0 | 0.001 | 0.001 | 0.001 |
| 59 | 10 | 9 | 0 | 0.000 | 0.000 | 0.000 |

n.risk (number at risk at time $t$ ), n.event (number of events at time $t$ ),
surv (Survival probability), UCI (upper confident interval limit),
LCI (lower confident interval limit).

**Table 14:**
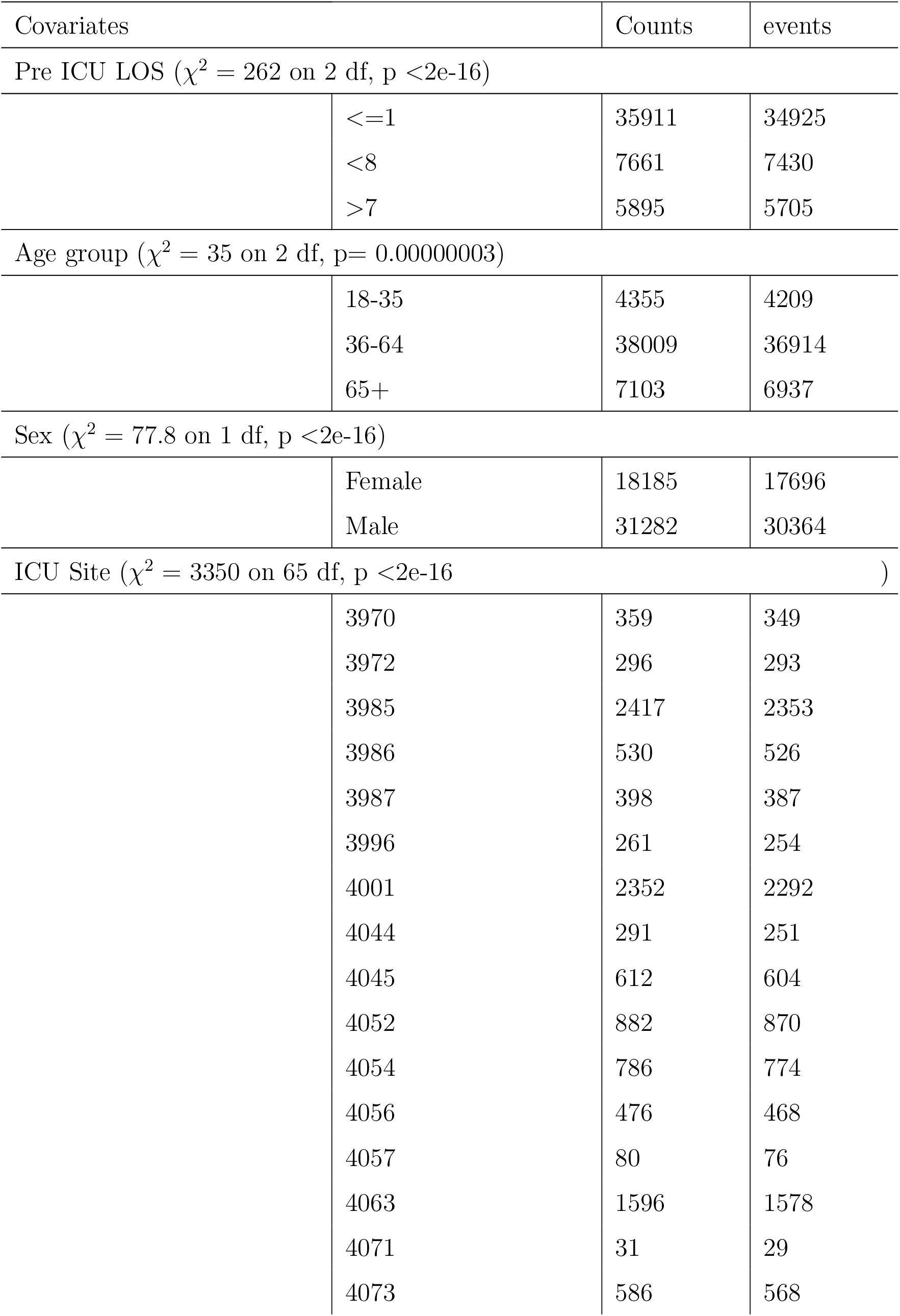
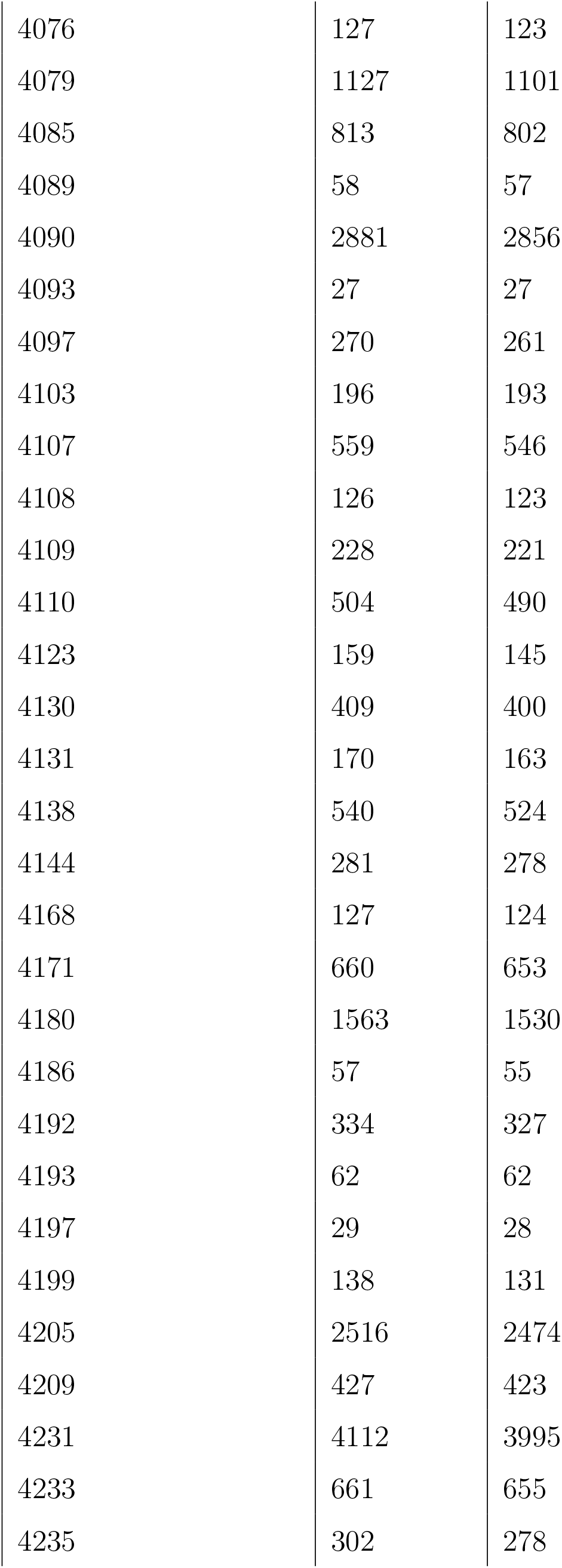
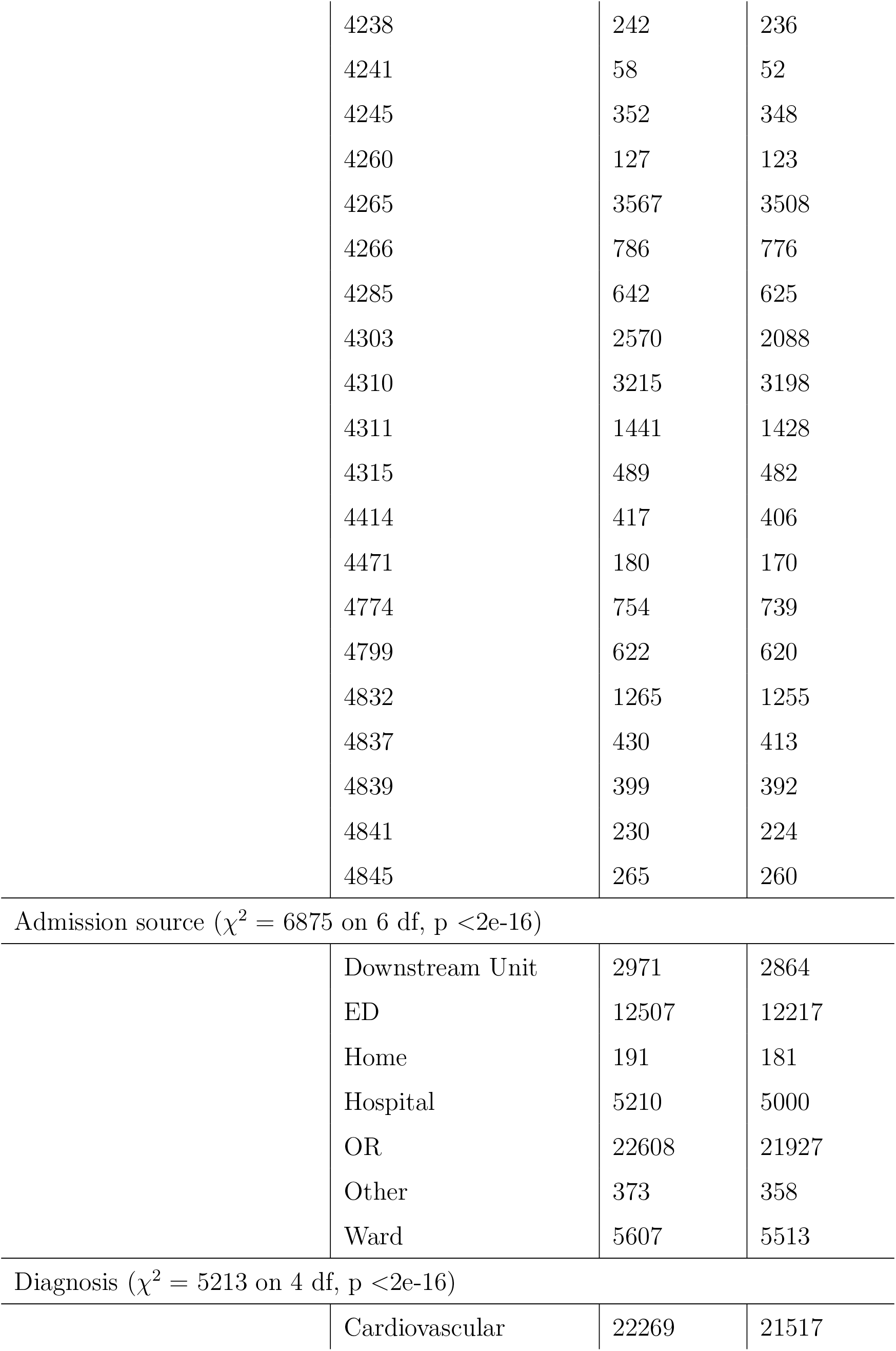
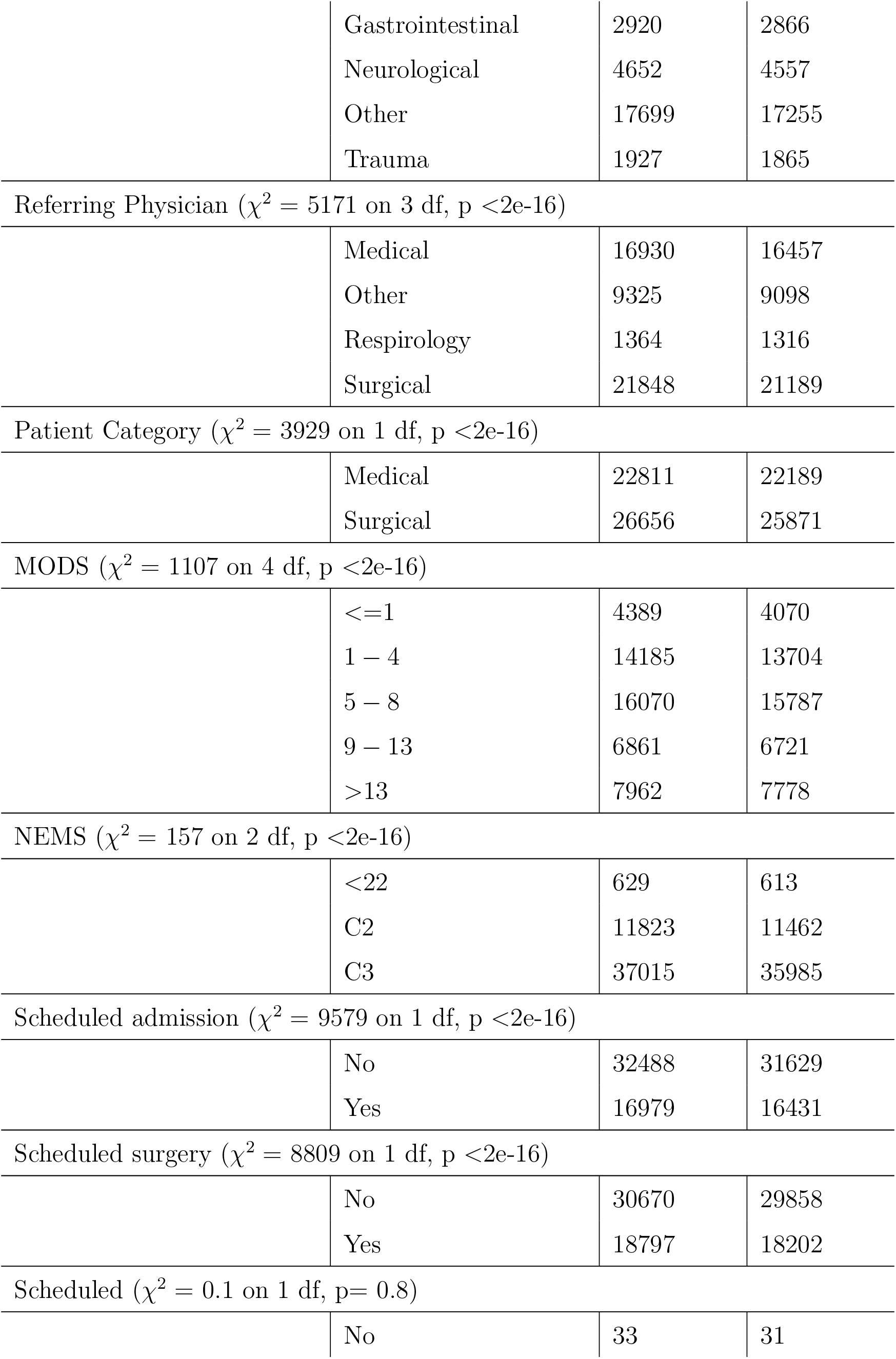
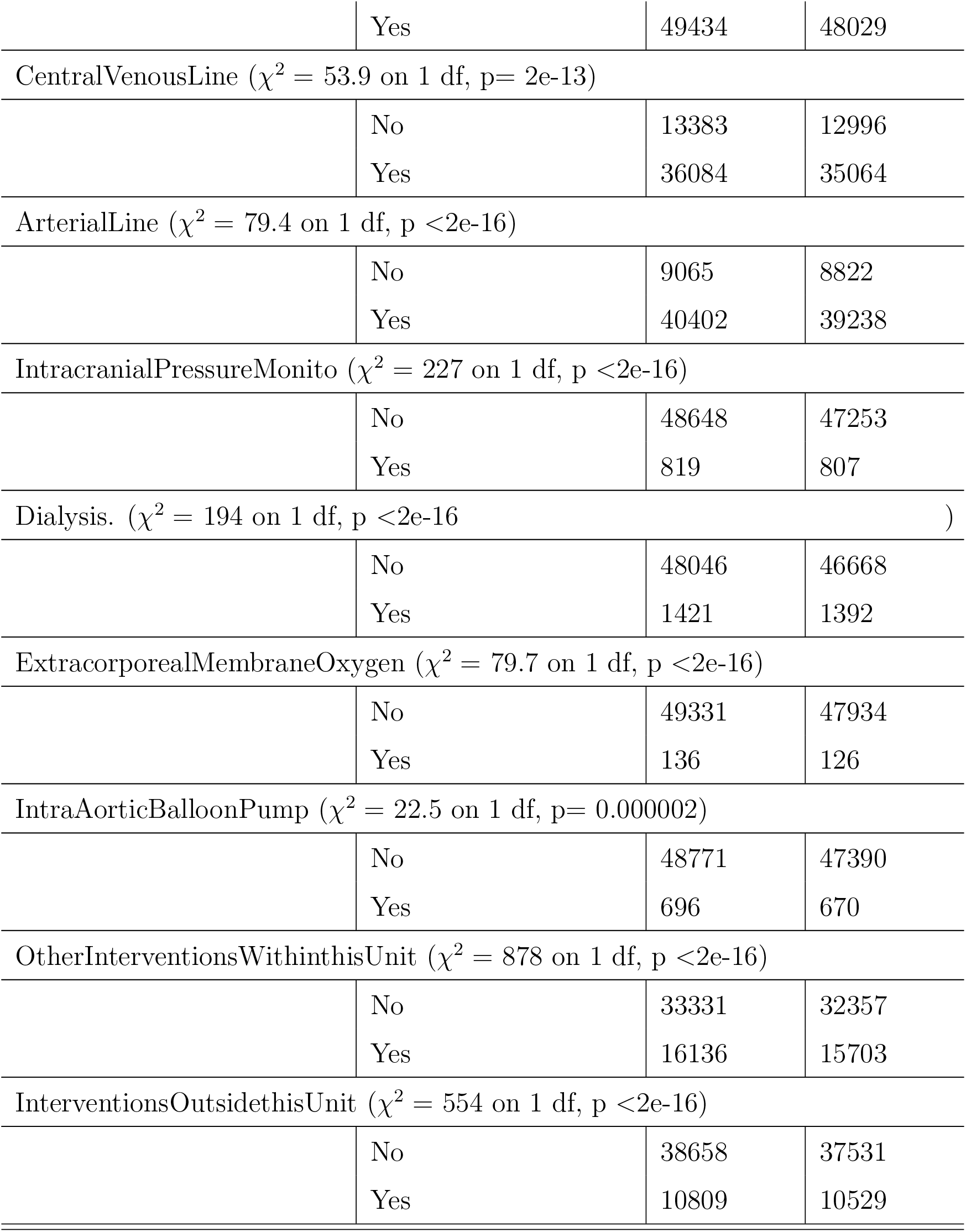
Description of the Log rank uni-variate test

| Covariates |  | Counts | events |
| --- | --- | --- | --- |
| Pre ICU LOS ($\chi^2 = 262$ on 2 df, $p < 2e-16$) | | | |
| | $\leq 1$ | 35911 | 34925 |
| | $< 8$ | 7661 | 7430 |
| | $> 7$ | 5895 | 5705 |
| Age group ($\chi^2 = 35$ on 2 df, $p = 0.00000003$) | | | |
|  | 18-35 | 4355 | 4209 |
|  | 36-64 | 38009 | 36914 |
|  | 65+ | 7103 | 6937 |
| Sex ($\chi^2 = 77.8$ on 1 df, $p < 2e-16$) | | | |
|  | Female | 18185 | 17696 |
|  | Male | 31282 | 30364 |
| ICU Site ($\chi^2 = 3350$ on 65 df, $p < 2e-16$) | | | |
|  | 3970 | 359 | 349 |
|  | 3972 | 296 | 293 |
|  | 3985 | 2417 | 2353 |
|  | 3986 | 530 | 526 |
|  | 3987 | 398 | 387 |
|  | 3996 | 261 | 254 |
|  | 4001 | 2352 | 2292 |
|  | 4044 | 291 | 251 |
|  | 4045 | 612 | 604 |
|  | 4052 | 882 | 870 |
|  | 4054 | 786 | 774 |
|  | 4056 | 476 | 468 |
|  | 4057 | 80 | 76 |
|  | 4063 | 1596 | 1578 |
|  | 4071 | 31 | 29 |
|  | 4073 | 586 | 568 |

|  |  |  |
| --- | --- | --- |
| 4076 | 127 | 123 |
| 4079 | 1127 | 1101 |
| 4085 | 813 | 802 |
| 4089 | 58 | 57 |
| 4090 | 2881 | 2856 |
| 4093 | 27 | 27 |
| 4097 | 270 | 261 |
| 4103 | 196 | 193 |
| 4107 | 559 | 546 |
| 4108 | 126 | 123 |
| 4109 | 228 | 221 |
| 4110 | 504 | 490 |
| 4123 | 159 | 145 |
| 4130 | 409 | 400 |
| 4131 | 170 | 163 |
| 4138 | 540 | 524 |
| 4144 | 281 | 278 |
| 4168 | 127 | 124 |
| 4171 | 660 | 653 |
| 4180 | 1563 | 1530 |
| 4186 | 57 | 55 |
| 4192 | 334 | 327 |
| 4193 | 62 | 62 |
| 4197 | 29 | 28 |
| 4199 | 138 | 131 |
| 4205 | 2516 | 2474 |
| 4209 | 427 | 423 |
| 4231 | 4112 | 3995 |
| 4233 | 661 | 655 |
| 4235 | 302 | 278 |
|  | 4238 | 242 | 236 |
|  | 4241 | 58 | 52 |
|  | 4245 | 352 | 348 |
|  | 4260 | 127 | 123 |
|  | 4265 | 3567 | 3508 |
|  | 4266 | 786 | 776 |
|  | 4285 | 642 | 625 |
|  | 4303 | 2570 | 2088 |
|  | 4310 | 3215 | 3198 |
|  | 4311 | 1441 | 1428 |
|  | 4315 | 489 | 482 |
|  | 4414 | 417 | 406 |
|  | 4471 | 180 | 170 |
|  | 4774 | 754 | 739 |
|  | 4799 | 622 | 620 |
|  | 4832 | 1265 | 1255 |
|  | 4837 | 430 | 413 |
|  | 4839 | 399 | 392 |
|  | 4841 | 230 | 224 |
|  | 4845 | 265 | 260 |
| Admission source ($\chi^2 = 6875$ on 6 df, p <2e-16) | | | |
|  | Downstream Unit | 2971 | 2864 |
|  | ED | 12507 | 12217 |
|  | Home | 191 | 181 |
|  | Hospital | 5210 | 5000 |
|  | OR | 22608 | 21927 |
|  | Other | 373 | 358 |
|  | Ward | 5607 | 5513 |
| Diagnosis ($\chi^2 = 5213$ on 4 df, p <2e-16) | | | |
|  | Cardiovascular | 22269 | 21517 |
|  | Gastrointestinal | 2920 | 2866 |
|  | Neurological | 4652 | 4557 |
|  | Other | 17699 | 17255 |
|  | Trauma | 1927 | 1865 |
| Referring Physician ($\chi^2 = 5171$ on 3 df, $p < 2e-16$) | | | |
|  | Medical | 16930 | 16457 |
|  | Other | 9325 | 9098 |
|  | Respirology | 1364 | 1316 |
|  | Surgical | 21848 | 21189 |
| Patient Category ($\chi^2 = 3929$ on 1 df, $p < 2e-16$) | | | |
|  | Medical | 22811 | 22189 |
|  | Surgical | 26656 | 25871 |
| MODS ($\chi^2 = 1107$ on 4 df, $p < 2e-16$) | | | |
| | $\leq 1$ | 4389 | 4070 |
|  | 1 – 4 | 14185 | 13704 |
|  | 5 – 8 | 16070 | 15787 |
|  | 9 – 13 | 6861 | 6721 |
| | $> 13$ | 7962 | 7778 |
| NEMS ($\chi^2 = 157$ on 2 df, $p < 2e-16$) | | | |
| | $< 22$ | 629 | 613 |
|  | C2 | 11823 | 11462 |
|  | C3 | 37015 | 35985 |
| Scheduled admission ($\chi^2 = 9579$ on 1 df, $p < 2e-16$) | | | |
|  | No | 32488 | 31629 |
|  | Yes | 16979 | 16431 |
| Scheduled surgery ($\chi^2 = 8809$ on 1 df, $p < 2e-16$) | | | |
|  | No | 30670 | 29858 |
|  | Yes | 18797 | 18202 |
| Scheduled ($\chi^2 = 0.1$ on 1 df, $p = 0.8$) | | | |
|  | No | 33 | 31 |
|  | Yes | 49434 | 48029 |
| CentralVenousLine ($\chi^2 = 53.9$ on 1 df, p= 2e-13) | | | |
|  | No | 13383 | 12996 |
|  | Yes | 36084 | 35064 |
| ArterialLine ($\chi^2 = 79.4$ on 1 df, p <2e-16) | | | |
|  | No | 9065 | 8822 |
|  | Yes | 40402 | 39238 |
| IntracranialPressureMonito ($\chi^2 = 227$ on 1 df, p <2e-16) | | | |
|  | No | 48648 | 47253 |
|  | Yes | 819 | 807 |
| Dialysis. ($\chi^2 = 194$ on 1 df, p <2e-16 ) | | | |
|  | No | 48046 | 46668 |
|  | Yes | 1421 | 1392 |
| ExtracorporealMembraneOxygen ($\chi^2 = 79.7$ on 1 df, p <2e-16) | | | |
|  | No | 49331 | 47934 |
|  | Yes | 136 | 126 |
| IntraAorticBalloonPump ($\chi^2 = 22.5$ on 1 df, p= 0.000002) | | | |
|  | No | 48771 | 47390 |
|  | Yes | 696 | 670 |
| OtherInterventionsWithinthisUnit ($\chi^2 = 878$ on 1 df, p <2e-16) | | | |
|  | No | 33331 | 32357 |
|  | Yes | 16136 | 15703 |
| InterventionsOutsidethisUnit ($\chi^2 = 554$ on 1 df, p <2e-16) | | | |
|  | No | 38658 | 37531 |
|  | Yes | 10809 | 10529 |

**Table 15:** Test of Cox PH assumption

| Covariate | $\chi^2$ | df | $p$-value |
| --- | --- | --- | --- |
| Site Code | 1156.267 | 65 | $< 2e - 16$ |
| Admission Source | 847.177 | 6 | $< 2e - 16$ |
| Diagnosis | 851.135 | 4 | $< 2e - 16$ |
| Scheduled ICU Admission | 953.906 | 1 | $< 2e - 16$ |
| Scheduled Surgery | 884.831 | 1 | $< 2e - 16$ |
| Patient Category | 609.514 | 1 | $< 2e - 16$ |
| Physician Specialty | 688.851 | 3 | $< 2e - 16$ |
| Central Venous Line | 29.441 | 1 | $5.8e - 08$ |
| Arterial Line | 0.206 | 1 | 0.6502 |
| Intracranial Pressure Monitor. | 69.925 | 1 | $< 2e - 16$ |
| Dialysis | 33.588 | 1 | $6.8e - 09$ |
| Extra-corporeal Membrane Oxygen | 7.091 | 1 | 0.0077 |
| Intra-Aortic Balloon Pump | 21.868 | 1 | $2.9e-06$ |
| Other Interventions Within this Unit | 113.692 | 1 | $< 2e - 16$ |
| Interventions Outside this Unit | 186.073 | 1 | $< 2e - 16$ |
| Age | 27.107 | 1 | $1.9e-07$ |
| Gender | 8.828 | 1 | 0.0030 |
| Age group | 7.381 | 2 | 0.0250 |
| Pre LOS | 28.340 | 2 | $7.0e-07$ |
| MODS Cat | 145.383 | 4 | $< 2e - 16$ |
| NEMS Cat | 21.775 | 2 | $1.9e-05$ |
| GLOBAL | 2552.177 | 101 | $< 2e - 16$ |
| $\chi^2$ (Chi-square value), df (degree of freedom) | | | |

**Table 16:** Survival distribution of log-logistic AFT model for CCIS data

| Covariate | est | L95% | U95% | exp(est) | L95% | U95% |
| --- | --- | --- | --- | --- | --- | --- |
| shape | 2.26 | 2.24 | 2.28 | NA | NA | NA |
| scale | 1.11 | 0.96 | 1.27 | NA | NA | NA |
| SiteCode 3972 | 0.31 | 0.16 | 0.46 | 1.36 | 1.18 | 1.58 |
| SiteCode 3985 | 0.39 | 0.28 | 0.50 | 1.48 | 1.32 | 1.64 |
| SiteCode 3986 | 0.48 | 0.34 | 0.61 | 1.61 | 1.41 | 1.84 |
| SiteCode 3987 | 0.22 | 0.08 | 0.36 | 1.25 | 1.09 | 1.43 |
| SiteCode 3996 | 0.55 | 0.39 | 0.71 | 1.73 | 1.48 | 2.03 |
| SiteCode 4001 | 0.20 | 0.09 | 0.31 | 1.22 | 1.09 | 1.36 |
| SiteCode 4044 | 0.34 | 0.18 | 0.50 | 1.40 | 1.20 | 1.64 |
| SiteCode 4045 | 0.25 | 0.12 | 0.37 | 1.28 | 1.13 | 1.45 |
| SiteCode 4052 | 0.35 | 0.23 | 0.47 | 1.41 | 1.25 | 1.59 |
| SiteCode 4054 | 0.43 | 0.31 | 0.55 | 1.53 | 1.36 | 1.73 |
| SiteCode 4056 | 0.49 | 0.35 | 0.62 | 1.62 | 1.42 | 1.86 |
| SiteCode 4057 | 0.10 | -0.12 | 0.33 | 1.11 | 0.89 | 1.39 |
| SiteCode 4063 | 0.03 | -0.08 | 0.14 | 1.03 | 0.92 | 1.15 |
| SiteCode 4071 | 0.07 | -0.25 | 0.40 | 1.08 | 0.78 | 1.49 |
| SiteCode 4073 | 0.31 | 0.19 | 0.44 | 1.36 | 1.20 | 1.55 |
| SiteCode 4076 | 0.57 | 0.36 | 0.78 | 1.76 | 1.43 | 2.18 |
| SiteCode 4079 | 0.43 | 0.31 | 0.55 | 1.54 | 1.37 | 1.73 |
| SiteCode 4085 | 0.40 | 0.28 | 0.52 | 1.49 | 1.32 | 1.68 |
| SiteCode 4089 | 0.44 | 0.17 | 0.70 | 1.55 | 1.19 | 2.02 |
| SiteCode 4090 | 0.27 | 0.16 | 0.38 | 1.31 | 1.18 | 1.46 |
| SiteCode 4093 | -0.29 | -0.64 | 0.06 | 0.75 | 0.53 | 1.06 |
| SiteCode 4097 | 0.41 | 0.25 | 0.56 | 1.50 | 1.29 | 1.75 |
| SiteCode 4103 | 0.19 | 0.03 | 0.35 | 1.21 | 1.03 | 1.42 |
| SiteCode 4107 | 0.46 | 0.33 | 0.59 | 1.58 | 1.39 | 1.80 |
| SiteCode 4108 | 0.71 | 0.51 | 0.91 | 2.04 | 1.67 | 2.49 |
| SiteCode 4109 | 0.26 | 0.10 | 0.42 | 1.30 | 1.11 | 1.53 |
| SiteCode 4110 | 0.11 | -0.01 | 0.24 | 1.12 | 0.99 | 1.27 |
| SiteCode 4123 | 0.30 | 0.13 | 0.47 | 1.35 | 1.13 | 1.60 |
| SiteCode 4130 | 0.37 | 0.23 | 0.51 | 1.45 | 1.26 | 1.66 |
| SiteCode 4131 | 0.04 | -0.14 | 0.21 | 1.04 | 0.87 | 1.24 |
| SiteCode 4138 | 0.63 | 0.50 | 0.77 | 1.88 | 1.65 | 2.15 |
| SiteCode 4144 | 0.47 | 0.31 | 0.62 | 1.59 | 1.37 | 1.86 |
| SiteCode 4168 | 0.22 | 0.03 | 0.42 | 1.25 | 1.03 | 1.52 |
| SiteCode 4171 | 0.43 | 0.31 | 0.56 | 1.54 | 1.36 | 1.75 |
| SiteCode 4180 | 0.17 | 0.06 | 0.28 | 1.19 | 1.06 | 1.33 |
| SiteCode 4186 | 0.26 | 0.00 | 0.52 | 1.30 | 1.00 | 1.69 |
| SiteCode 4192 | 0.51 | 0.37 | 0.65 | 1.66 | 1.44 | 1.92 |
| SiteCode 4193 | 0.35 | 0.08 | 0.62 | 1.42 | 1.09 | 1.86 |
| SiteCode 4197 | 0.05 | -0.32 | 0.42 | 1.06 | 0.73 | 1.53 |
| SiteCode 4199 | -0.12 | -0.30 | 0.06 | 0.89 | 0.74 | 1.07 |
| SiteCode 4205 | 0.47 | 0.36 | 0.58 | 1.60 | 1.43 | 1.78 |
| SiteCode 4209 | 0.15 | 0.01 | 0.28 | 1.16 | 1.01 | 1.33 |
| SiteCode 4231 | 0.21 | 0.10 | 0.32 | 1.23 | 1.11 | 1.37 |
| SiteCode 4233 | 0.28 | 0.15 | 0.40 | 1.32 | 1.16 | 1.49 |
| SiteCode 4235 | 0.33 | 0.17 | 0.48 | 1.39 | 1.19 | 1.62 |
| SiteCode 4238 | 0.37 | 0.21 | 0.52 | 1.44 | 1.24 | 1.68 |
| SiteCode 4241 | 0.22 | -0.05 | 0.49 | 1.25 | 0.96 | 1.63 |
| SiteCode 4245 | 0.28 | 0.14 | 0.42 | 1.33 | 1.15 | 1.53 |
| SiteCode 4260 | 0.29 | 0.07 | 0.50 | 1.33 | 1.08 | 1.65 |
| SiteCode 4265 | 0.58 | 0.47 | 0.69 | 1.79 | 1.60 | 1.99 |
| SiteCode 4266 | 0.55 | 0.43 | 0.68 | 1.74 | 1.54 | 1.96 |
| SiteCode 4285 | 0.19 | 0.07 | 0.32 | 1.21 | 1.07 | 1.37 |
| SiteCode 4303 | 0.49 | 0.38 | 0.60 | 1.63 | 1.46 | 1.82 |
| SiteCode 4310 | 0.29 | 0.18 | 0.39 | 1.33 | 1.20 | 1.48 |
| SiteCode 4311 | 0.26 | 0.15 | 0.37 | 1.30 | 1.16 | 1.45 |
| SiteCode 4315 | 0.37 | 0.24 | 0.50 | 1.45 | 1.27 | 1.66 |
| SiteCode 4414 | 0.37 | 0.22 | 0.52 | 1.44 | 1.25 | 1.67 |
| SiteCode 4471 | 0.28 | 0.11 | 0.45 | 1.32 | 1.11 | 1.57 |
| SiteCode 4774 | 0.35 | 0.23 | 0.48 | 1.42 | 1.26 | 1.61 |
| SiteCode 4799 | 0.46 | 0.34 | 0.59 | 1.59 | 1.40 | 1.81 |
| SiteCode 4832 | 0.33 | 0.22 | 0.45 | 1.40 | 1.24 | 1.57 |
| SiteCode 4837 | 0.44 | 0.30 | 0.58 | 1.55 | 1.35 | 1.79 |
| SiteCode 4839 | 0.73 | 0.58 | 0.88 | 2.08 | 1.79 | 2.41 |
| SiteCode 4841 | 0.44 | 0.27 | 0.60 | 1.55 | 1.31 | 1.83 |
| SiteCode 4845 | 0.47 | 0.31 | 0.62 | 1.60 | 1.36 | 1.87 |
| Admission Source ED | -0.13 | -0.17 | -0.08 | 0.88 | 0.84 | 0.92 |
| Admission Source Home | 0.13 | -0.01 | 0.27 | 1.14 | 0.99 | 1.31 |
| Admission Source Hospital | 0.11 | 0.06 | 0.16 | 1.12 | 1.07 | 1.17 |
| Admission Source OR | -0.31 | -0.36 | -0.26 | 0.73 | 0.70 | 0.77 |
| Admission Source Other | 0.15 | 0.04 | 0.26 | 1.17 | 1.04 | 1.30 |
| Admission Source Ward | -0.13 | -0.18 | -0.08 | 0.88 | 0.84 | 0.92 |
| Diagnosis Gastrointestinal | 0.28 | 0.24 | 0.33 | 1.33 | 1.27 | 1.38 |
| Diagnosis Neurological | 0.31 | 0.27 | 0.35 | 1.37 | 1.31 | 1.42 |
| Diagnosis Other | 0.40 | 0.37 | 0.43 | 1.49 | 1.45 | 1.53 |
| Diagnosis Trauma | 0.52 | 0.47 | 0.58 | 1.69 | 1.59 | 1.79 |
| Scheduled ICU Admission Yes | -0.22 | -0.27 | -0.17 | 0.80 | 0.77 | 0.85 |
| Scheduled Surgery Yes | -0.16 | -0.21 | -0.11 | 0.85 | 0.81 | 0.90 |
| Patient Category Surgical | 0.01 | -0.04 | 0.06 | 1.01 | 0.96 | 1.06 |
| Physician Specialty Other | -0.01 | -0.04 | 0.03 | 0.99 | 0.96 | 1.03 |
| Physician Specialty Respiriology | 0.15 | 0.09 | 0.22 | 1.16 | 1.09 | 1.24 |
| Physician Specialty Surgical | -0.08 | -0.13 | -0.03 | 0.92 | 0.88 | 0.97 |
| Central Venous Line Yes | 0.17 | 0.14 | 0.19 | 1.18 | 1.16 | 1.21 |
| Arterial Line Yes | 0.19 | 0.16 | 0.21 | 1.20 | 1.17 | 1.24 |
| Intracranial Pressure Monitor Yes | 0.55 | 0.48 | 0.63 | 1.74 | 1.61 | 1.87 |
| Dialysis Yes | 0.01 | -0.04 | 0.07 | 1.01 | 0.96 | 1.07 |
| Extra corporeal Membrane Oxygen Yes | 0.67 | 0.49 | 0.85 | 1.95 | 1.63 | 2.33 |
| Intra Aortic Balloon Pump Yes | 0.33 | 0.25 | 0.40 | 1.39 | 1.29 | 1.50 |
| Other Interventions in this Unit Yes | 0.07 | 0.05 | 0.09 | 1.07 | 1.05 | 1.10 |
| Interventions Outside this Unit Yes | 0.02 | 0.00 | 0.04 | 1.02 | 1.00 | 1.04 |
| Gender Male | 0.00 | -0.01 | 0.02 | 1.00 | 0.99 | 1.02 |
| Age group2 | 0.12 | 0.08 | 0.16 | 1.13 | 1.08 | 1.18 |
| Age group3 | 0.05 | -0.01 | 0.12 | 1.05 | 0.99 | 1.12 |
| Pre LOSC2 | 0.04 | 0.02 | 0.06 | 1.04 | 1.02 | 1.07 |
| Pre LOSC3 | 0.15 | 0.12 | 0.18 | 1.16 | 1.12 | 1.19 |
| MODS CatC2 | 0.11 | 0.07 | 0.14 | 1.11 | 1.08 | 1.15 |
| MODS CatC3 | 0.22 | 0.19 | 0.26 | 1.25 | 1.21 | 1.29 |
| MODS CatC4 | 0.27 | 0.23 | 0.31 | 1.31 | 1.26 | 1.36 |
| MODS CatC5 | 0.18 | 0.14 | 0.21 | 1.19 | 1.15 | 1.24 |
| NEMS CatC2 | -0.01 | -0.09 | 0.06 | 0.99 | 0.91 | 1.06 |
| NEMS CatC3 | 0.04 | -0.04 | 0.11 | 1.04 | 0.96 | 1.12 |

**Table 17:** Variable selection criteria for models fitted to the data using backward elimination.

| Variable removed | df | Deviance | Pr(>Chi) |
| --- | --- | --- | --- |
| Site Code | 65 | 1267.19 | 2.80E-222 |
| Admission Source | 6 | 452.51 | 1.42E-94 |
| Diagnosis | 4 | 889.69 | 2.86E-191 |
| ScheduledICUAdmission | 1 | 75.50 | 3.65E-18 |
| ScheduledSurgery | 1 | 38.93 | 4.38E-10 |
| PatientCategory (PC) | 1 | 0.10 | 0.7498 |
| PC + PhysicianSpecialty | 4 | 44.34 | 5.46E-09 |
| PC + CentralVenousLine | 2 | 184.81 | 7.40E-41 |
| PC + ArterialLine | 2 | 193.28 | 1.07E-42 |
| PC + IntracranialPressureMonitor | 2 | 210.01 | 2.50E-46 |
| PC + Dialysis (D) | 2 | 0.40 | 0.8200 |
| PC + D + ExtracorporealMembraneOxygen | 3 | 51.27 | 4.28E-11 |
| PC + D + IntraAorticBalloonPump | 3 | 77.06 | 1.31E-16 |
| PC + D + OtherInterventionsWithinthisUnit | 3 | 42.75 | 2.78E-09 |
| PC + D + Interventions Outside (IO) | 3 | 3.06 | 0.3826 |
| PC + D + IO + Gender | 4 | 3.24 | 0.5179 |
| PC + D + IO + Gender + Age_group | 6 | 77.82 | 1.00E-14 |
| PC + D + IO + Gender + Pre_LOS | 6 | 100.58 | 1.90E-19 |
| PC + D + IO + Gender + MODS | 8 | 273.48 | 1.79E-54 |
| PC + D + IO + Gender + NEMS | 6 | 26.86 | 0.0002 |

